# Reopening California : Seeking Robust, Non-Dominated COVID-19 Exit Strategies

**DOI:** 10.1101/2021.04.26.21256105

**Authors:** Pedro Nascimento de Lima, Robert Lempert, Raffaele Vardavas, Lawrence Baker, Jeanne Ringel, Carolyn M. Rutter, Jonathan Ozik, Nicholson Collier

**Affiliations:** RAND Corporation, Santa Monica, CA 90401, USA; Pardee RAND Graduate School, Santa Monica, CA 90401, USA; Argonne National Laboratory, Lemont, IL 60439, USA

## Abstract

Amid global scarcity of COVID-19 vaccines and the threat of new variant strains, California and other jurisdictions face the question of when and how to implement and relax COVID-19 Nonpharmaceutical Interventions (NPIs). While policymakers have attempted to balance the health and economic impacts of the pandemic, decentralized decision-making, deep uncertainty, and the lack of widespread use of comprehensive decision support methods can lead to the choice of fragile or inefficient strategies. This paper uses simulation models and the Robust Decision Making (RDM) approach to stress-test California’s reopening strategy and other alternatives over a wide range of futures. We find that plans which respond aggressively to initial outbreaks are required to robustly control the pandemic. Further, the best plans adapt to changing circumstances, lowering their stringent requirements to reopen over time or as more constituents are vaccinated. While we use California as an example, our results are particularly relevant for jurisdictions where vaccination roll-out has been slower.

## 1 Introduction

The COVID-19 pandemic has wreaked havoc across the US, causing over 30 million confirmed cases and 500,000 related deaths, and state-wide shutdowns of all non-essential businesses. As state officials navigate the pandemic, they must manage both health and economic goals. Generally, reopening plans [1–4] have been proposed as phased strategies, in which the state allows economic activity to resume based on meeting COVID-19 incidence targets. However, pandemics often follow oscillatory waves of infection, and many states have already been forced to revise their plans, either shutting down after new outbreaks or adjusting which activities are allowed in each phase. To ensure long-term success in combating the pandemic, local policymakers need to consider the trade-offs underlying reopening decisions, while accounting for deep uncertainties.

Since one-time lockdowns have proven to be insufficient to control the pandemic, a coherent, long-term strategy is needed. Instead of adopting a stable, pre-defined strategy, local policymakers have changed regulations and instated NPIs adaptively, often adopting NPIs based on the decisions of other jurisdictions [5]. A key challenge in recommending a stable, transparent strategy - i.e., a clear prescription of which NPIs local policymakers should enact given a set of observed conditions - is to account for the effects of the biological, behavioral, and technological uncertainties surrounding the pandemic. One year into the pandemic, many factors and constraints are unknown and outside of the control of policymakers, including vaccine efficacy to prevent transmission [6], the behavioral response to vaccination (i.e., change in population mixing behaviors after vaccination) and vaccine uptake. Variant strains with higher transmissibility [7] might hamper the effects of social distancing measures and might impact vaccine efficacy.

Model-based analyses of alternative strategies for defending society against the COVID-19 pandemic have been invaluable. Prior studies evaluating how local governments should manage NPIs in the absence of COVID-19 vaccines have presented a grim outlook [8–11] justifying the need for stringent NPI strategies, and have revealed the impracticality of strategies based on naturally-acquired herd immunity [12]. Analyses specifically focusing on vaccine strategies accounting for age-dependent mortality and vaccine scarcity [13–16] have generally supported ACIP’s vaccine allocation recommendations [17] if one seeks to minimize deaths. More recently, analyses focusing on how to relax NPIs in the presence of vaccination generally find that premature reopening would result in resurgences and potentially compromise the benefits of vaccination [18–23].

This paper offers three contributions to the emerging literature of NPI strategies in the presence of COVID-19 vaccines. First, we evaluate a range of alternatives to manage NPI levels, including strategies that resemble reopening plans with fixed thresholds such as California’s Blueprint for a Safe economy [3] and CDC’s Operational Strategy for Reopening Schools [24] as well as alternative strategies that change those thresholds over time. Second, we consider economic consequences of NPIs, allowing the analysis of robustness trade-offs among strategies. Third, we employ the Robust Decision Making (RDM) approach [25–27] to facilitate comparison of alternative adaptive NPI strategies and to address the deep uncertainties [28] surrounding the COVID-19 pandemic and vaccination rollout. Although RDM has proven useful in other policy areas where uncertainties abound (i.e., climate change [29], coastal resilience [30], terrorism insurance [31], water resources management [32]), this approach has been underutilized in the public health policy literature. This approach allow us to seek and find robust, non-dominated reopening strategies. Using the state of California as an example, we demonstrate that strategies with fixed thresholds can be pareto-dominated^1^ by strategies with time-varying or endogenous thresholds. This analysis and approach might support changes in reopening plans in the wake of vaccination roll-out.

The paper is organized as follows. First, the methods section provides a high-level overview of our approach, and details on the model can be found in our prior work [10, 33] and in the Supplemental Information section. Once the problem framing is defined, we focus on the main substantive finding that strategies with fixed thresholds might be dominated by strategies with time-varying thresholds. Finally, we discuss how improved strategies might be implemented, as well as potential challenges in pursuing pareto-efficient strategies.

## 2 Methods

### 2.1 Problem Framing

State public health departments need to decide how to manage their NPI levels using a coherent set of rules that seek to minimize both the health and economic impacts of the pandemic. California’s Blueprint for a Safer Economy plan [3], for example, requires the number of daily new cases to be below 7 cases / 100 k and positivity rates to be below 8 % to allow counties to move below their most stringent NPI level (widespread). As of Feb 1st, 2021, 99% of the state population was living under the widespread risk level, in which some non-essential businesses are closed or operate under restrictions. We frame the decision problem as to how to define the threshold criteria in those plans over time seeking to balance health and other welfare goals.

Reopening plans are not only defined by threshold criteria but also contain a larger set of decisions not analyzed in this paper. These decisions include which businesses are allowed to operate under each risk level, capacity constraints, and a set of adaptation measures. These decisions directly affect societal welfare by imposing differential costs on specific activities and have evolved over time within California’s reopening plan [3]. While examining these decisions could reveal that targeted interventions have the potential to produce pareto-improvements [34], doing so is beyond the scope of this paper. Therefore, this paper holds the definition of the risk levels constant and asks *how* one should navigate between the risk levels over time.

Although states can mandate low-cost mitigation measures such as voluntary social distancing and mask-wearing that have proven effective [35], imperfect compliance still allows the virus to circulate among the population in many US states. Therefore, the key decision-making challenge this paper investigates is how to manage blunt NPIs (such as restricting economic activity or interrupting in-person education), contingent on the net effect of other low-cost measures such as mask-wearing, behavioral changes, and physical adaptation measures. High compliance to low-cost measures and initial lockdowns, followed by tight strict movement interventions (e.g, New Zealand’s strategy), and appropriate levels of testing, contact tracing, and isolation to prevent re-seeding could potentially eliminate COVID-19 locally and obviate the need for this decision-making process and reopening plans. Reality has shown, however, that US states and many countries have failed to control the spread of the virus through those instruments, thus forcing local decision-makers to still face trade-offs.

When crafting reopening plans, decision-makers should seek strategies that are *both robust and non-dominated* - that is, they not only perform well across a wide range of futures (robustness), but they also do not make society unnecessarily worse-off with respect to a set of relevant outcomes (pareto-efficiency)^2^. Because pareto-dominated policies make members of society unnecessarily worse-off, seeking pareto-efficient policies is an essential goal of policy analysis. While seeking robust, non-dominated policies, decision-makers have to consider a number of factors, including the goals that one seeks to achieve, the policy levers that are within reach, uncertainties that can influence the decision between those policy levers, and how those elements are connected. Table 1 contains the main elements of this decision problem that are included in our analysis, using an XLRM framework [26]. The letters X, L, R, and M refer to four categories of factors important to the analysis: outcome measures (M) that reflect decision makers’ goals; policy levers (L) that decision-makers use to pursue their goals; uncertainties (X) that may affect the connection between levers and outcomes; and relationships (R), instantiated in the simulation model, that link uncertainties and levers to outcomes. The subsequent sections provide details on each of those four basic elements.

**Table 1:** XLRM framework - uncertainties, policy levers, relationships and metrics. The XLRM framework provides an overview of the relevant components in an RDM analysis. Each component in the list of policy levers is used to define a set of strategies. In this paper, we explore 78 strategies, which are defined in Appendix I.

| X - Uncertainties | L - Policy levers |
| --- | --- |
| • Vaccine efficacy to prevent transmission<br>• Loss of immunity<br>• Behavioral response to vaccination<br>• Willingness to vaccinate<br>• Changes in transmissibility (i.e., induced by variant strains)<br>• Actual vaccination Rate | • Baseline level of caution $x_b$<br>• NPI strategy $s \in \{C, T, V\}$<br>• Time-based strategies $s = T$<br>– Level of caution factor $\alpha$<br>– Transition date $T_\alpha$<br>• Vaccination-based strategies $s = V$<br>– Vaccination reference point $V_{mid}$<br>– Relaxation rate $k_c$ |
| R - Relationships (models) | M - Metrics |
| Meta-population deterministic ODE [10, 33] | 75<sup>th</sup> Regret percentile of deaths / 100 k people, years of life lost, cases, income loss, and days under NPIs |
| Computable general equilibrium model [36] |  |

### 2.2 Relationships

This paper builds from the models underlying RANDs COVID-19 State Policy Tool [10, 33, 36]. The original model and tool use a state-level deterministic epidemiological model [33] and a general equilibrium economic model [36] to inform state policymakers on the trade-offs of alternative NPIs. The set of NPIs used by each US state is characterized by a discrete set of intervention levels ranging from 1 (no intervention) to 6 (close schools, bars, restaurants, and nonessential businesses; and issue a shelter-in-place order for everyone but essential workers). The economic model [36] provides an estimate of weekly income loss for each US state and each intervention level, which we integrate over time based on the NPI levels used in the epidemiological model. The epidemiological model contains five strata: those below 18 years (Young), those with more than 65 years with or without chronic conditions, frontline essential workers, working-age individuals with chronic conditions, and working-age individuals without chronic conditions. Each intervention level is associated with population mixing matrices that describe how strata interact with each other in six different settings: household, work, school, commercial, recreation, and other [10, 33]. Interventions are modeled as changing the level of mixing which occurs in each of these settings. For instance, closing schools reduces school and work mixing but increases home mixing. Given the specified model structure, the NPI time series, and the mixing matrices, we can calibrate and run our model for any US state using cumulative monthly deaths time series [37]. In this analysis, the model was calibrated in the period of March 1st 2020 through Dec 25th 2020 for the state of California. The policies are evaluated from Dec 25th through Jan 31st, 2021. Further details about our model can be found in the Supplemental Information (SI) section.

### 2.3 Policy Levers and Strategies

Following the definitions commonly used in state-level plans, we define NPIs as alternative sets of intervention levels. Within each NPI level, a set of restrictions are imposed. For instance, when *NPI*_*t*_ = 1, the lowest level, businesses and schools are opened, although society may still have inexpensive policies in place, such as mask-wearing. When *NPI*_*t*_ = 6, the highest level, society imposes the most stringent restrictions on people leaving their homes and interacting with others.

An NPI Strategy consists of a rule that determines how NPI levels change over time. The time-dependent NPI level can be specified as a function of COVID-19 prevalence *p*_*t*_ using the controller function:

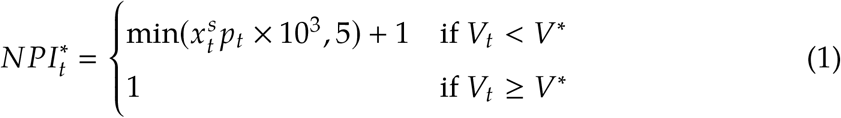

where *p*_*t*_ represents estimated COVID-19 prevalence scaled by 10^3^, and 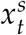 represents the sensitivity of policymakers to this prevalence which we label “level of caution”, *V*_*t*_ is the current vaccination coverage rate, and *V*^∗^ is the threshold vaccination coverage rate at which policymakers terminate the use of NPI’s. The controller function is bound between one and six, corresponding to the six intervention levels in our model. This results in a policy wherein a 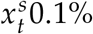 increase in COVID-19 prevalence results in policymakers increasing restriction levels by one level until the highest restriction level is adopted. This controller function provides a simple representation of the types of phased reopening plans used by US states, such as California’s Blueprint for a Safer Economy plan [3]. Eq. 1 also reflects the fact that policy-makers might exhibit different levels of sensitivity to changes in COVID-19 prevalence, and that this sensitivity 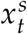 can change over time.

In this study, we ask how 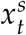 should be managed over time. We evaluate three types of NPI Strategies, denoted by *s* = *C, T*, and *V*, which differ in how the level of caution 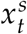 is managed. The strategies can use a constant level of caution (*s* = *C*), a two-step function of time (*s* = *T*), or a smooth function of the proportion of the population that is vaccinated (*s* = *V*). We define each type of strategy as follows:

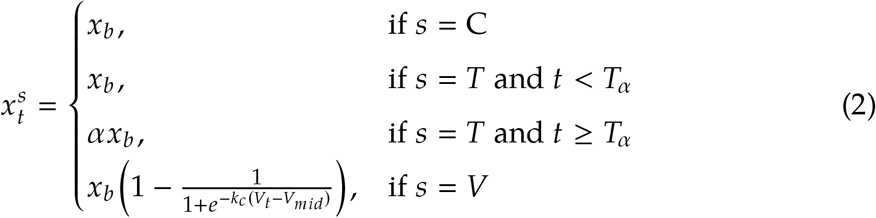

The constant level of caution strategy (*s* = *C*) holds 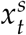 constant at a baseline level of caution *x*_*b*_. The two-step strategy time-based strategy (*s* = *T*) begins with the value *x*_*b*_ and changes to *αx*_*b*_ at a predefined date *T*_*α*_ (i.e., *T*_*α*_ could be the beginning of spring, or the fall). We assume, that and 0 > *α* > 1. The vaccination-based strategy (*s* = *V*) calculates a time-varying level of caution as a smooth function of the cumulative number of persons fully vaccinated *V*_*t*_ using an inverse logistic function that starts at 1 when *V*_*t*_ = 0, and ends at zero when *V*_*t*_ = 1, with mid-point *V*_*mid*_, and curvature defined by *kc*. A particular NPI strategy is defined by the choice of values for the parameters *s, x*_*b*_, *α, V*_*mid*_, and *k*_*c*_.

Figure 1 illustrates the model dynamics with a constant level of caution *x*_*t*_. This formulation leads to frequent interventions, similar to patterns seen in California. Note that with a low level of caution (*x*_*b*_ = 0.5) prevalence is allowed to increase substantially before a state takes action. With a high level of caution (*x*_*b*_ = 24) prevalence is held much lower. As a reference, California’s current strategy is represented by a level of caution of *x*_*t*_ ≈ 5.^3^

**Figure 1:**
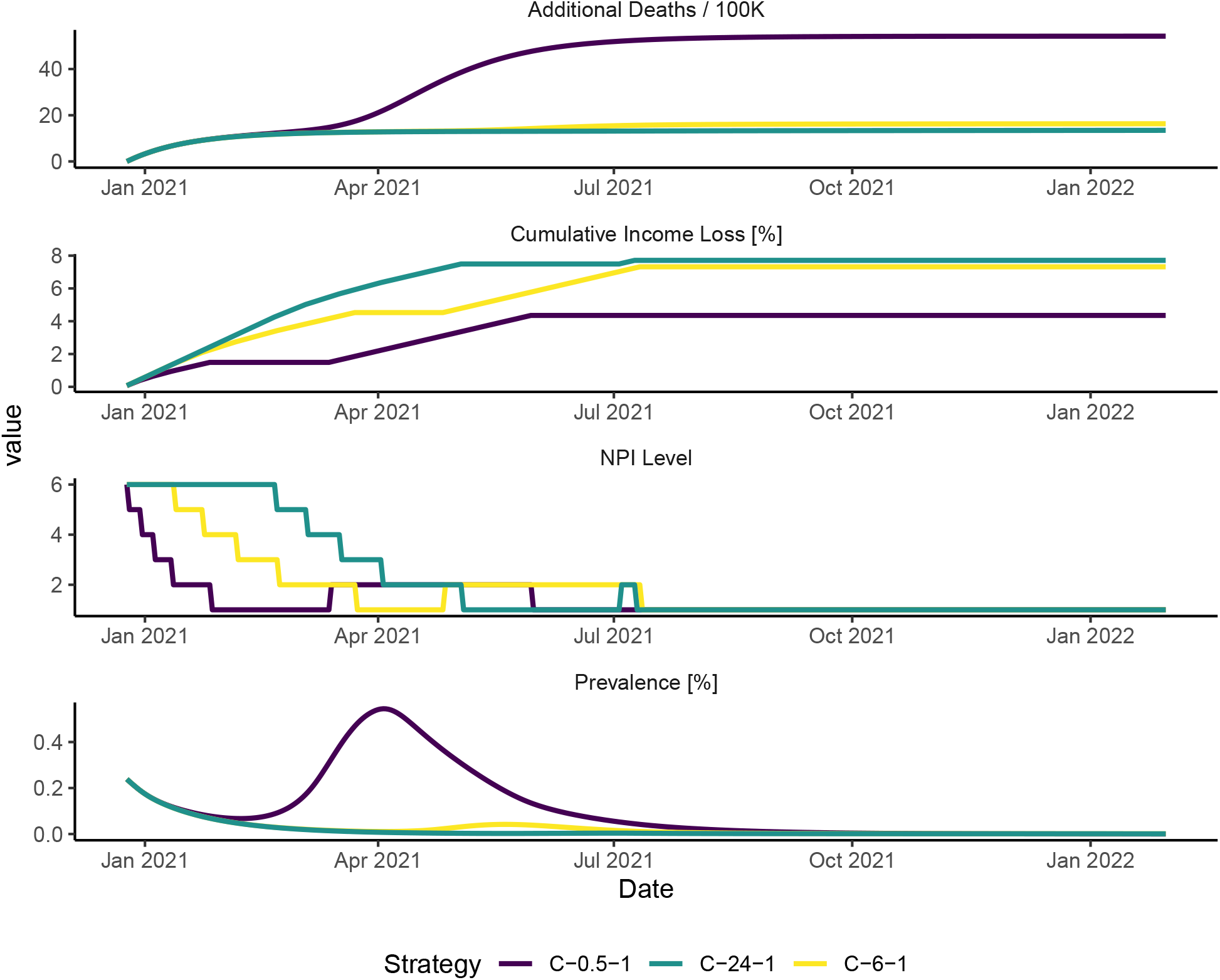
Model dynamics with adaptive NPI strategies in California. This figure presents three illustrative runs testing three strategies in an example future state of the world. Strategies are coded as follows throughout this paper. The first letter in the strategy code represents the strategy type (C for constant caution, T for time-based, and V for vaccine-based strategies). The subsequent number represents the baseline level of caution *x*_*b*_, and the third number is a sequential code to make the strategy code unique. The full list of strategies tested in this paper is provided in Appendix I. In this figure, the most stringent strategy, C-24-1 uses a level of caution *x*_*t*_ = 24 throughout the simulation run. Strategy C-6-1 uses a level of caution *x*_*t*_ = 6 and produces similar results, while strategy C-0.5-1 results in a substantial increase in deaths. None of these model runs represent a forecast.

Eqs 1 and 2 represent a range of alternative strategies to manage NPIs during vaccination roll-out. If California were to change its thresholds and reduce restrictions in the fall of 2021, that could be represented by a time-based strategy *s* = *T* with a baseline level of caution *x*_*b*_ = 5, transition time *T*_*α*_ = *Fall*, 2021 and a reduction factor of *α* = 0.5. If *α* = 0, California would allow businesses and in-person education to operate at *T*_*α*_. Similarly, California might decide to slowly scale down restrictions based on the number of vaccinations, such that restrictions would be halved when 50% of the population is vaccinated. This policy would be represented by a strategy *s* = *V*, with *x*_*b*_ = 5 and *V*_*mid*_ = 0.5. We consider a set of strategies by combining strategy types and their parameters in a full-factorial experimental design. The full list of strategies and the parameters used in this paper is provided in Appendix I.

### 2.4 Uncertainties

We categorize uncertainties into two distinct classes: well-characterized uncertainties and deep uncertainties. Well-characterized uncertainties are those for which historical data or clinical evidence can provide information, either directly or through calibration. Deep uncertainties [28] are those for which calibration or existing clinical evidence provides little information at the time of this writing, and have a high potential to affect the choice of the strategies.

Well-characterized uncertainties include the length of disease states, which are set to fixed values based on published findings, and parameters that are selected using model calibration, including the magnitude of the seasonal effect on mixing. Calibrated parameters are chosen using the Incremental Mixture Approximate Bayesian Computation (IMABC) approach [39]. The IMABC algorithm results in 100 simulated draws from the posterior distribution of the parameter set. This parameter set, hereafter termed “calibrated parameters”, contains values for 42 model parameters, including information about how effective NPIs have been in the past, mortality rates, and disease progression rates.

Deep uncertainties include vaccine efficacy to prevent transmission, the behavioral mixing response to vaccination, willingness to vaccinate, changes in transmissibility, immunity duration, and the actual vaccination rate. Uncertainties surrounding vaccine efficacy are particularly concerning given their impact on the pandemic dynamics. While vaccine efficacy to prevent disease has been established, vaccine efficacy to prevent infection is unknown at the time of this writing [6, 40, 41]. Similarly, the effect of new variant strains on transmissibility and other disease parameters is of particular concern. For example, variants B.1.1.7, B.1.351, and P.1 have demonstrated an impact on transmissibility, inactivity, and antibody escape capabilities[7].

To examine the impact of deep uncertainties on future outcomes, we first draw a quasi-random sample of 200 unknown parameter vectors using Latin Hypercube sampling to ensure representation of the parameter space. Details about these parameters and their bounds are provided in Supplemental Materials under the Experimental Design section. We then combine each row in our “calibrated parameters” dataset with each row in our Latin Hypercube to create a new dataset with 20,000 rows in which each row represents a single future state of the world (SOW). The final experimental design is obtained by testing each strategy in each of the 20,000 future states of the world. We define our set of strategies using a full-factorial experimental design yielding 78 strategies, described in Appendix I. Hence, our experimental design contains 1.56 million cases (78 strategies * 20,000 future states of the world). Following the RDM approach [25], we evaluate how each strategy would perform in a wide range of futures, and judge those strategies by their ability to cope with many potential futures.

### 2.5 Outcome Measures

When judging alternative reopening strategies, policymakers often have to weigh multiple criteria to make decisions. Our epidemiological model computes the cumulative number of COVID-19 cases per 100,000 people including undetected cases, years of life lost due to COVID-19 deaths per 100,000 people, and COVID-19 deaths per 100,000 people. Reopening decisions, however, can have far-reaching social welfare consequences which are not explicitly computed in COVID-19 epidemiological models. While short-term economic impacts might have been limited in some circumstances [42], the full social welfare cost of NPIs includes the effects of interrupted in-person education [43], mental health impacts of isolation, other illness exacerbated by reduced use of non-COVID health services, impacts of financial effects on mental and physical health, deaths of despair [44], and long-term loss of income. While we are not aware of a published estimation of the total welfare loss induced by NPIs, one needs to find criteria that can be computed from the model and are proportional to the marginal welfare loss induced by alternative NPI levels.

One criterion one might use is the number of days under NPIs (*NPI*_*t*_ > 1), which, in our model, is equivalent to the number of days with school closures. This is the outcome used in this paper to assess pareto-efficiency. While it is easily interpreted, this number does not distinguish scenarios in which non-essential businesses are closed for long periods, so other proxies for welfare loss might be desirable. One approach is to use weights for each NPI level, such that those weights are proportional to the marginal daily welfare loss induced by each NPI level. With the purpose of demonstrating how this could be done, we use an estimate of Income Loss [36] as those weights to present a proxy for economic consequences of NPI restrictions. Although these proxies are imperfect measures of social welfare loss induced by NPIs, our conclusions do not rely on their precision, but on the assumption that NPI costs are increasing in the level of restriction. This structural assumption allows us to illuminate trade-offs and reveal pareto-dominated strategies. Therefore, our findings do not rely on the precision of any welfare loss estimate, but on the structure of the epidemiological model^4^.

Because our interest lies in the robustness of strategies rather than in their optimality for any particular future, we use Regret [25] as a robustness metric and the 75th Regret percentile as a decision criterion. Regret is computed for each metric of interest as follows. We construct a dataset of model runs in which each row contains the values of the outcomes defined above at the end of the simulation. Each row in this dataset represents the performance of each strategy on each future state of the world (SOW) characterized by uncertainties as described previously. Robustness in this study is operationalized with a separate regret metric for each outcome of interest. Regret *R*_*s, f*_ is defined for each strategy *s* in each SOW *f* as the difference between the observed outcome and the best possible outcome in that future:

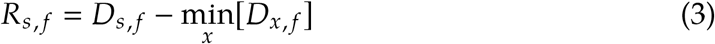

The goal of the decision-making process is to adopt a strategy that minimizes regret across a wide range of potential futures, across all of the outcomes of interest. When trying to minimize regret across a wide range of outcomes and futures, decision-makers often find that prioritizing a single set of outcomes imposes regret on other outcomes. Similarly, minimizing regret in a single future might create vulnerabilities to other future states of the world. While a thorough discussion on the use of regret metrics is beyond the scope of this paper, the interested reader can refer to methodological pieces discussing the use of robustness metrics [25, 26].

## 3 Results

This section describes two main results from our analysis, namely: i) the trade-offs among the baseline and alternative strategies, and ii) the existence and characteristics of dominated policies. While traditional decision analyses often rule out dominated policies as a first step, we find it useful to focus our discussion on those strategies with the goal of illuminating potential alternatives.

Figure 2 (A) presents the distribution of Deaths / 100,000 people for each strategy, which are defined in Table 2. These results emphasize that only strategies that use a high baseline level of caution are able to control deaths and thus result in low regret in terms of deaths. The strategies using lower baseline levels of caution unsurprisingly result in higher death regret. These results demonstrate that the baseline level of caution non-linearly affects deaths. For example, figure 2 (B) demonstrates that there is a significant reduction in the number of deaths when one compares the level of caution 0.5 to 1.5, but using a level of caution of 24 instead of 12 marginally reduces deaths.

**Table 2:** 75^*th*^ Regret percentiles by metric and strategy for a set of non-dominated and constant-caution strategies. The table presents a subset of the strategies evaluated in this paper including only Non-Dominated strategies and Constant strategies. The complete list of strategies is available in Appendix I. The first letter in the strategy code stands for the strategy type (C for constant caution, T for time-based, and V for vaccine-based). The subsequent number represents the baseline level of caution *x*_*b*_, and the third number is a sequential number making the strategy code unique. The parameters column describes policy levers that characterize each strategy, as described in the methods section. The final three columns present the 75^*th*^ regret percentile of three metrics of interest.

| Strategy | Strategy Parameters | Deaths / 100k | NPI Days |
| --- | --- | --- | --- |
| C-0.5-1 | constant | 93 | 73 |
| C-1.5-1 | constant | 50 | 161 |
| C-3-1 | constant | 28 | 209 |
| C-6-1 | constant | 12 | 238 |
| C-12-1 | constant | 4 | 247 |
| C-24-1 | constant | 0 | 252 |
| V-24-2 | $V^* = 60\%; k_c = 15$ | 1 | 248 |
| V-24-6 | $V^* = 50\%; k_c = 15$ | 3 | 240 |
| V-24-5 | $V^* = 50\%; k_c = 10$ | 2 | 244 |
| T-24-4 | $\alpha = 50\%; T_\alpha = 2021 - 03 - 10$ | 4 | 229 |
| T-24-3 | $\alpha = 10\%; T_\alpha = 2021 - 09 - 26$ | 5 | 225 |
| T-12-2 | $\alpha = 10\%; T_\alpha = 2021 - 07 - 04$ | 18 | 192 |
| T-24-2 | $\alpha = 10\%; T_\alpha = 2021 - 07 - 04$ | 9 | 213 |

**Figure 2:**
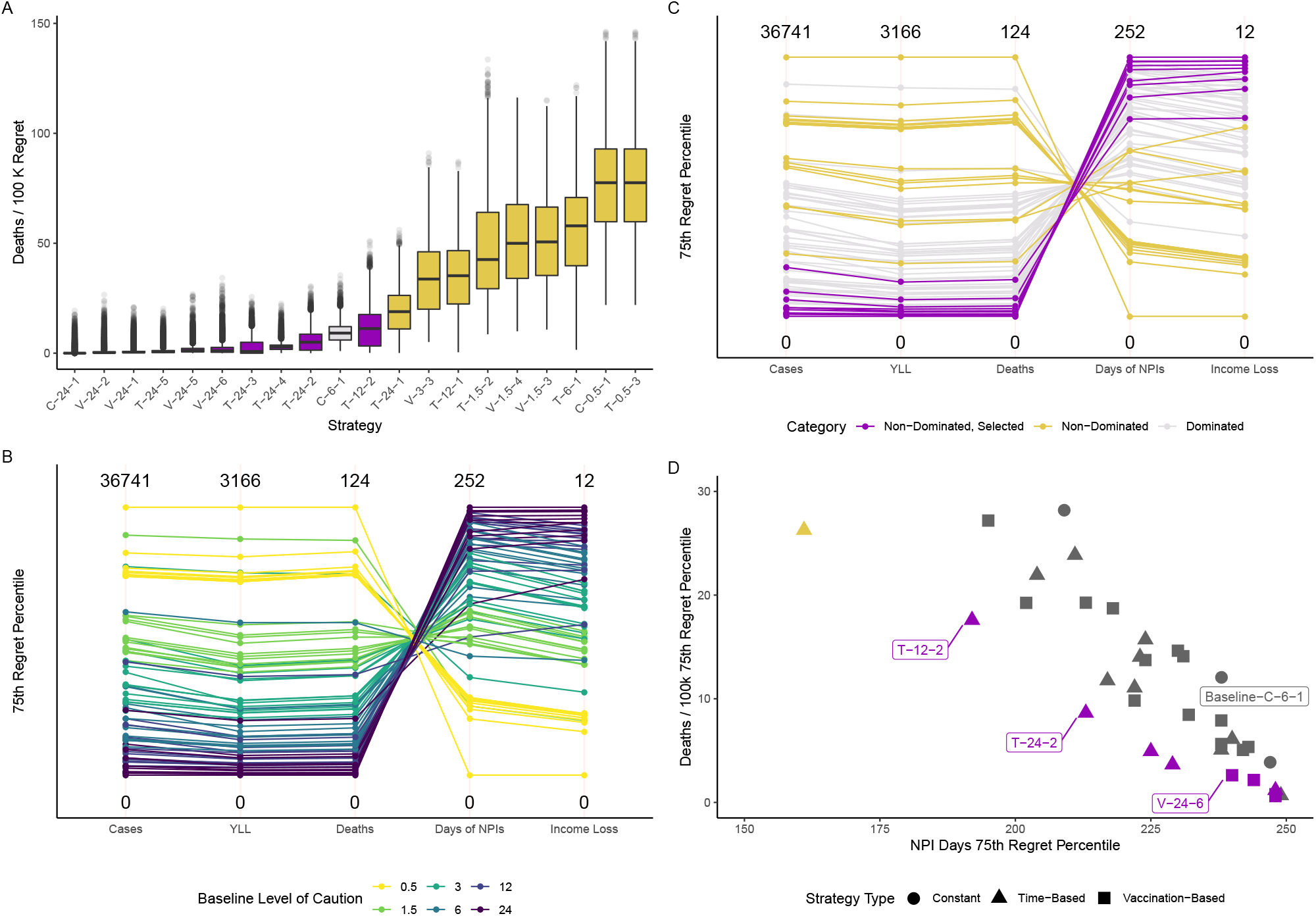
Robustness Trade-offs emerging from 78 alternative reopening strategies. Panel A shows the distribution of a Regret metric for four outcomes of interest. Each line in Panel B and C represents a single strategy. Vertical axes represent the 75^*th*^ Regret percentile for each metric. All Health outcomes are normalized per 100 thousand people. Strategies are coded as follows. The first letter indicates whether the NPI strategy uses a Time-based level of caution (T), a Vaccination-Based level of caution (V), or a Constant level of caution (C). The subsequent number describes the baseline level of caution *x*_*b*_ and a third number is a sequential number that creates a unique code for each strategy. Table 2 contains a description of all strategies.

The baseline strategy with a fixed level of caution (C-6-1) seems to achieve a compromise when compared to more stringent strategies. More stringent strategies (i.e., C-24-1, C-12-1) are able to achieve a lower number of deaths regret, but doubling the level of caution does not halve the number of deaths, nor does it double the social welfare costs, as measured by Loss of Income or Days under NPIs. This result indicates that a baseline strategy with a level of caution *x*_*b*_ > 6 is necessary to robustly control the number of deaths. If the goal of policymakers was only to minimize the number of deaths from the pandemic, these results would imply that the best approach would be to sustain the highest level of caution indefinitely.

Preventing deaths, however, comes with a cost. Figure 2 (B) contains other outcomes produced by the set of strategies evaluated. In figure 2 B, each line represents one of the 78 tested strategies, and the parallel axes summarize the performance of each strategy using the 75^*th*^percentile ^5^ of the regret distribution shown in panel A for each one of the metrics. Again, this figure reveals that only strategies with higher baseline levels of caution (i.e., *x*_*b*_ > 3) are able to reach the bottom of the Deaths regret scale. Unsurprisingly, strategies that achieve the lowest numbers of deaths are also the ones that generate the highest social welfare costs, as measured by the two proxies available. Although the order of magnitude of the outcomes can vary among other models, this finding is in line with prior studies using multiple models [8].

Although the baseline strategy (C-6-1) seems to produce acceptable health outcomes given the other alternatives, that does not imply that this strategy is pareto-efficient. To assess pareto-efficiency, we categorize strategies into three classes. Non-dominated strategies are those that are not outperformed by any other strategy with respect to health and social welfare outcomes using the 75^*th*^ regret percentile as a criterion^6^. We select strategies with relatively low numbers of deaths seeking to represent a decision-maker that is willing to prioritize health outcomes. Finally, dominated strategies are those that exhibit worse health or social welfare outcomes. These strategies are color-coded in panels C and D of figure 2.

Figure 2 (D) reveals a troubling pattern for strategies based on constant thresholds. The figure illustrates that strategies with constant levels of caution (circles) are dominated by *many* vaccination and time-based strategies. Vaccination-based strategies (squares) and time-based strategies (triangles) are closer to the origin in that plot. We find that strategies using a constant level of caution (the circles in figure 2 D) are systematically dominated, except for the most stringent strategy or the most relaxed strategies. That is to say, unless society is willing to indefinitely sustain the highest level of stringency we considered, sustaining a fixed level of caution is always dominated by strategies that change the level of caution over time.

The strategies that dominate the baseline strategy (C-6-1) while resulting in a lower or equivalent number of deaths share the same characteristics. They start with a higher level of caution, then relax as vaccination and time advances. For example, strategy T-24-2 starts with a higher level of caution of 24 and relaxes its level of caution by 90% on the fourth of July. This strategy outperforms the baseline strategy by achieving lower social welfare and health regrets. Starting with high stringency levels then relaxing when immunity is widespread is arguably what other countries with higher capacity to control COVID-19 (i.e., New Zealand) will do and our results support that strategy. Although California’s Blueprint for a Safer economy plan originally had fixed thresholds, California has imposed a regional stay-at-home order from December 2020 through January 2021, effectively increasing the level of caution in that period. Recently, California updated its plan and made it dependent on vaccination rollout. Our results generally support changes on reopening plans that effectively make reopening policies more stringent while immunity is not widespread. Nonetheless, our results also demonstrate that stress-testing a wide range of strategies against other alternatives is important to ensure that the selected policies are not pareto-dominated by other alternatives.

While simpler SEIR models with homogenous populations might produce similar results, that does not need to be the case. We attribute our results and the rationale be-hind the vaccine-based strategies to the interactions between the vaccination strategy being used and the heterogeneities included in our model. As more individuals from the most vulnerable groups are immunized first, the average infection-fatality ratio (IFR) amongst the currently infected should decrease, thus suggesting that strategies with a fixed level of caution would be dominated. In the limit, as we approach an endemic state [45], the marginal benefit of NPIs decrease and the strategies accounting for these dynamics dominate strategies with fixed thresholds. The time scale of that transition depends on several factors, including heterogeneous fatality rates, vaccination strategy, and the rate at which immunity increases in the population. Other models, such as similar ODEs models or agent-based models that incorporate those heterogeneities should reach similar conclusions.

## 4 Conclusion

Our main substantive conclusion is that adaptive reopening strategies with fixed thresholds can be dominated by alternatives that are more stringent but change their stringency over time. While this finding points to potentially better policies, it also demonstrates that seemingly sensible reopening policies might not be pareto-efficient. This finding has important implications for existing reopening plans, not only in California but also for other US states and countries pursuing similar reopening strategies. These findings suggest that localities with stringent policies might have to craft time or vaccine-based reopening policies as vaccination is made widely available. Similarly, jurisdictions pursuing strategies with a low baseline level of caution might be trading death regret for small benefits, and are vulnerable to the emergence of new, more transmissible strains. Failing to cautiously adjust reopening policies will result in excess deaths from premature reopening decisions and/or unnecessary economic burden on those most vulnerable. While balancing multiple outcomes has not generally been done formally (i.e., we fail to find published rigorous analyses comprising a wide range of alternatives associated with reopening plans), these decisions are arguably the most important policy decisions of 2021.

After demonstrating that current strategies can be pareto-dominated, the next question is whether and how pareto-efficient strategies can be implemented. This paper does not offer a fixed timetable for when thresholds should be changed because doing so would defeat the purpose of our quest for robust reopening strategies. In our model, strategies are defined with respect to local conditions such as the number of cases and vaccination rates, which may be different across regions. However, after running our analysis and considering a large experimental design for a single state as we did for California, one can find a time-based reopening strategy that approximates the performance of the robust vaccination-based strategy, provided that vaccination progresses as expected. Translating a vaccination-based strategy to a time-based might prove useful for implementation purposes because vaccination strategies were defined as smooth functions of vaccination. Because vaccination rates are not equal across jurisdictions and the immunity status of the population would differ, the resulting timetable would be different for each jurisdiction, but would likely dominate alternative fixed-threshold strategies. Nevertheless, the general result that fixed-threshold reopening policies are dominated would still hold because they are a result of model structure, not parameters that describe the population of California.

The disparities exacerbated by the pandemic offer a compelling reason for more concern and rigor in defining reopening plans. While affluent populations are hedged against health and economic risks by savings and remote employment, vulnerable populations - within the US and abroad - have been experiencing the worst of the pandemic. Vulnerable populations were more likely to have had COVID-19 [34], be denied in-person education [43], and experience hunger during the pandemic [46]. How NPIs are managed in the next several months will determine the outcomes of the COVID-19 pandemic and will shape the trade-offs that these populations face. While our paper did not explicitly evaluate outcomes for distinct sub-populations within California or other jurisdictions, the evidence so far overwhelmingly points to the conclusion that populations at the margins are likely to pay a high proportion of the costs presented in this analysis, and therefore will bear the burden of dominated reopening strategies. If policymakers choose pareto-dominated reopening strategies, then it is inevitable that the already vulnerable populations will be the most affected by the consequences of dominated decisions.

Even after accounting for multiple uncertainties and simulating a wide range of strategies under many conditions, our analysis still presents limitations. While we account for three behavioral responses in our model (increase in mixing due to vaccination, change in transmissibility driven by changes in behaviors, and willingness to vaccinate), this analysis does not contain an endogenous behavioral response to changing prevalence other than the effects induced by the NPIs. In reality, people likely voluntarily react to COVID-19 prevalence, businesses adapt their operations to reduce the risk of transmission, and all these responses endogenously reduce the need for state-mandated NPIs. Similarly, when individuals or businesses relax, new surges can happen. These endogenous behavioral responses would likely introduce additional oscillations and dynamic challenges to the NPIs and might even dominate model dynamics if included. However, we are not aware of comprehensive behavioral models that we could confidently apply to our model at this stage. Better incorporating plausible behavioral mechanisms in our model is one of the next steps in our research agenda, and could reveal even more interesting results.

Although we consider immunity duration as an uncertainty, our analysis does not explicitly account for multiple components of immunological protection that will likely influence the transition to an endemic state [45]. However, doing so would likely strengthen the case for time-varying strategies. If sterilizing immunity is short-lived but second infections have a substantially lower IFR, then loss of immunity will represent a smaller challenge going forward, thus requiring a lower level of intervention in the future. This analysis also does not address other long-term outcomes from alternative policies (lack of traditional education in the long-term educational outcomes, long-term COVID-19 health effects such as lung damage, mental health impacts of isolation, other illness exacerbated by reduced use of non-COVID health services, the impact of financial effects on mental and physical health, etc.).

Another limitation is a result of our model structure. The model used in this analysis is a deterministic ODE model with heterogeneous population strata, implying that eradicating COVID-19 is never achieved and that the state. While this assumption might be reasonable for US states given the lack of coordination among states, this assumption is not reasonable for smaller countries or countries with tight travel controls that prevent re-seeding (e.g., New Zealand). Such countries can benefit from our framing but should adopt models that can represent eradication. Because our model is defined at the state level, this analysis also does not represent interactions among different geographic levels. Accounting for multiple geographic levels and re-seeding would also likely weaken the case for constant strategies and strengthen our conclusions. Finally, this analysis does not explicitly consider distributional concerns. While these limitations have the potential to shift the tradeoff curves, they are unlikely to change our substantive results. Future iterations of this analysis might choose to include these additional mechanisms and further stress test more policies against an even wider set of uncertainties.

Despite these limitations, this analysis demonstrated that the RDM approach can be useful to stress-test a wide range of COVID-19 reopening policies under conditions of deep uncertainty. More broadly, other decision-making under deep uncertainty (DMDU) methods and tools [28] might also prove useful. Our approach can accommodate the relaxation of the structural assumptions mentioned above, allowing the policy set to be tested in an increasingly larger experimental design, helping to meet the demand for rational policy-making during a pandemic [48]. In that regard, our work contributes to a stream of analyses [49, 50] and initiatives [51] that seek to address structural uncertainties in infectious disease models. RDM shares many of the goals of these approaches [50] but differs in how it addresses uncertainty. Understanding those differences and how DMDU methods can contribute to and learn from existing approaches used by infectious disease modelers might be useful to help policymakers make better decisions in this and the next pandemic.

## Data Availability

The code and data used in our analysis are available under a GPL-2.0 license.

https://github.com/RANDCorporation/covid-19-reopening-california

## Acknowledgements

We thank all our sponsors for their support. This research was funded by Mala Gaonkar and Surgo Foundation UK Limited, a separate legal entity to Surgo Ventures and the Anne and James Rothenberg Dissertation Award. The funding for the prior project to develop the epidemiological model was provided by gifts from RAND supporters and income from operations. We also thank the National Institute of Allergies and Infectious Diseases (R01AI118705) for providing support in projects that led to preliminary work and ideas that motivated this project. High-Performance Computing resources were provided by Laboratory Computing Resource Center at Argonne National Laboratory (Bebop cluster). Computing and labor resources provided by Argonne were funded by the U.S. Department of Energy, Office of Science, under contract number DE-AC02-06CH11357.

## Appendix I List of Strategies

Table 3 characterizes each strategy assessed in this paper. Each row represents one strategy, their corresponding parameters, and the 75^*th*^ percentile of the regret distribution over the 20,000 futures under which each strategy was tested. Lower regret represents better performance. The parameters in this table as used in the equations discussed in the Methods - Policy Levers section. Figure 3 illustrates that the information provided in table 3 is a summary of the regret distributions of each one of the strategies. While table 3 provides a summary of our findings, figure 3 illustrates how different strategies handle the challenges imposed by the uncertainties we used for our stress-tests, and how outcomes vary even when strategies are fixed. Figure 3 A shows that strategies with lower levels of caution tend to result in more deaths and lower numbers of days under NPIs. Strategies with higher levels of caution absorb those challenges by imposing longer intervention periods. Figure 3 B shows that adaptive strategies with different initial levels of caution can achieve similar results, hinting that it is possible to start with higher levels of caution and adaptively decrease the level of caution (e.g, T-12-2, T-24-2) while arriving at outcomes that are similar to outcomes obtained by strategies with constant levels of caution (e.g., C-6-1).

**Table 3:** Strategy characteristics and many-objective robustness measures.

| Strategy | Strategy Parameters | Deaths / 100k | NPI Days |
| --- | --- | --- | --- |
| C-0.5-1 | constant | 93 | 73 |
| C-1.5-1 | constant | 50 | 161 |
| C-3-1 | constant | 28 | 209 |
| C-6-1 | constant | 12 | 238 |
| C-12-1 | constant | 4 | 247 |

Table 3 – continued
| Strategy | Strategy Parameters | Deaths / 100k | NPI Days |
| --- | --- | --- | --- |
| C-24-1 | constant | 0 | 252 |
| V-0.5-2 | $V^* = 60\%; k_c = 15$ | 93 | 72 |
| V-1.5-2 | $V^* = 60\%; k_c = 15$ | 53 | 153 |
| V-3-2 | $V^* = 60\%; k_c = 15$ | 31 | 200 |
| V-6-2 | $V^* = 60\%; k_c = 15$ | 14 | 231 |
| V-12-2 | $V^* = 60\%; k_c = 15$ | 5 | 242 |
| V-24-2 | $V^* = 60\%; k_c = 15$ | 1 | 248 |
| V-0.5-1 | $V^* = 60\%; k_c = 10$ | 94 | 71 |
| V-1.5-1 | $V^* = 60\%; k_c = 10$ | 54 | 149 |
| V-3-1 | $V^* = 60\%; k_c = 10$ | 32 | 197 |
| V-6-1 | $V^* = 60\%; k_c = 10$ | 15 | 230 |
| V-12-1 | $V^* = 60\%; k_c = 10$ | 5 | 243 |
| V-24-1 | $V^* = 60\%; k_c = 10$ | 1 | 248 |
| V-0.5-6 | $V^* = 50\%; k_c = 15$ | 94 | 70 |
| V-1.5-6 | $V^* = 50\%; k_c = 15$ | 58 | 140 |
| V-3-6 | $V^* = 50\%; k_c = 15$ | 37 | 181 |
| V-6-6 | $V^* = 50\%; k_c = 15$ | 19 | 213 |
| V-12-6 | $V^* = 50\%; k_c = 15$ | 8 | 232 |
| V-24-6 | $V^* = 50\%; k_c = 15$ | 3 | 240 |
| V-0.5-5 | $V^* = 50\%; k_c = 10$ | 95 | 68 |
| V-1.5-5 | $V^* = 50\%; k_c = 10$ | 59 | 138 |
| V-3-5 | $V^* = 50\%; k_c = 10$ | 37 | 182 |
| V-6-5 | $V^* = 50\%; k_c = 10$ | 19 | 218 |
| V-12-5 | $V^* = 50\%; k_c = 10$ | 8 | 238 |
| V-24-5 | $V^* = 50\%; k_c = 10$ | 2 | 244 |
| V-0.5-4 | $V^* = 40\%; k_c = 15$ | 95 | 65 |
| V-1.5-4 | $V^* = 40\%; k_c = 15$ | 68 | 123 |
| V-3-4 | $V^* = 40\%; k_c = 15$ | 50 | 154 |
| V-6-4 | $V^* = 40\%; k_c = 15$ | 33 | 177 |
| V-12-4 | $V^* = 40\%; k_c = 15$ | 19 | 202 |
| V-24-4 | $V^* = 40\%; k_c = 15$ | 10 | 222 |
| V-0.5-3 | $V^* = 40\%; k_c = 10$ | 97 | 62 |

**Table 3 – continued**
| Strategy | Strategy Parameters | Deaths / 100k | NPI Days |
| --- | --- | --- | --- |
| V-1.5-3 | $V^* = 40\%; k_c = 10$ | 66 | 124 |
| V-3-3 | $V^* = 40\%; k_c = 10$ | 46 | 161 |
| V-6-3 | $V^* = 40\%; k_c = 10$ | 27 | 195 |
| V-12-3 | $V^* = 40\%; k_c = 10$ | 14 | 224 |
| V-24-3 | $V^* = 40\%; k_c = 10$ | 6 | 238 |
| T-0.5-6 | $\alpha = 50\%; T_\alpha = 2021 - 09 - 26$ | 93 | 73 |
| T-1.5-6 | $\alpha = 50\%; T_\alpha = 2021 - 09 - 26$ | 53 | 155 |
| T-3-6 | $\alpha = 50\%; T_\alpha = 2021 - 09 - 26$ | 31 | 200 |
| T-6-6 | $\alpha = 50\%; T_\alpha = 2021 - 09 - 26$ | 14 | 223 |
| T-12-6 | $\alpha = 50\%; T_\alpha = 2021 - 09 - 26$ | 5 | 238 |
| T-24-6 | $\alpha = 50\%; T_\alpha = 2021 - 09 - 26$ | 1 | 249 |
| T-0.5-5 | $\alpha = 50\%; T_\alpha = 2021 - 07 - 04$ | 93 | 72 |
| T-1.5-5 | $\alpha = 50\%; T_\alpha = 2021 - 07 - 04$ | 56 | 140 |
| T-3-5 | $\alpha = 50\%; T_\alpha = 2021 - 07 - 04$ | 34 | 191 |
| T-6-5 | $\alpha = 50\%; T_\alpha = 2021 - 07 - 04$ | 16 | 224 |
| T-12-5 | $\alpha = 50\%; T_\alpha = 2021 - 07 - 04$ | 6 | 240 |
| T-24-5 | $\alpha = 50\%; T_\alpha = 2021 - 07 - 04$ | 1 | 248 |
| T-0.5-4 | $\alpha = 50\%; T_\alpha = 2021 - 03 - 10$ | 104 | 53 |
| T-1.5-4 | $\alpha = 50\%; T_\alpha = 2021 - 03 - 10$ | 71 | 127 |
| T-3-4 | $\alpha = 50\%; T_\alpha = 2021 - 03 - 10$ | 46 | 165 |
| T-6-4 | $\alpha = 50\%; T_\alpha = 2021 - 03 - 10$ | 24 | 211 |
| T-12-4 | $\alpha = 50\%; T_\alpha = 2021 - 03 - 10$ | 11 | 222 |
| T-24-4 | $\alpha = 50\%; T_\alpha = 2021 - 03 - 10$ | 4 | 229 |
| T-0.5-3 | $\alpha = 10\%; T_\alpha = 2021 - 09 - 26$ | 93 | 73 |
| T-1.5-3 | $\alpha = 10\%; T_\alpha = 2021 - 09 - 26$ | 53 | 149 |
| T-3-3 | $\alpha = 10\%; T_\alpha = 2021 - 09 - 26$ | 36 | 182 |
| T-6-3 | $\alpha = 10\%; T_\alpha = 2021 - 09 - 26$ | 22 | 204 |
| T-12-3 | $\alpha = 10\%; T_\alpha = 2021 - 09 - 26$ | 12 | 217 |
| T-24-3 | $\alpha = 10\%; T_\alpha = 2021 - 09 - 26$ | 5 | 225 |
| T-0.5-2 | $\alpha = 10\%; T_\alpha = 2021 - 07 - 04$ | 93 | 72 |
| T-1.5-2 | $\alpha = 10\%; T_\alpha = 2021 - 07 - 04$ | 64 | 130 |
| T-3-2 | $\alpha = 10\%; T_\alpha = 2021 - 07 - 04$ | 54 | 147 |

| **Strategy** | **Strategy Parameters** | **Deaths / 100k** | **NPI Days** |
| --- | --- | --- | --- |
| T-6-2 | $\alpha = 10\%; T_\alpha = 2021 - 07 - 04$ | 33 | 168 |
| T-12-2 | $\alpha = 10\%; T_\alpha = 2021 - 07 - 04$ | 18 | 192 |
| T-24-2 | $\alpha = 10\%; T_\alpha = 2021 - 07 - 04$ | 9 | 213 |
| T-0.5-1 | $\alpha = 10\%; T_\alpha = 2021 - 03 - 10$ | 124 | 0 |
| T-1.5-1 | $\alpha = 10\%; T_\alpha = 2021 - 03 - 10$ | 109 | 65 |
| T-3-1 | $\alpha = 10\%; T_\alpha = 2021 - 03 - 10$ | 93 | 92 |
| T-6-1 | $\alpha = 10\%; T_\alpha = 2021 - 03 - 10$ | 71 | 112 |
| T-12-1 | $\alpha = 10\%; T_\alpha = 2021 - 03 - 10$ | 47 | 130 |
| T-24-1 | $\alpha = 10\%; T_\alpha = 2021 - 03 - 10$ | 26 | 161 |

**Figure 3:**
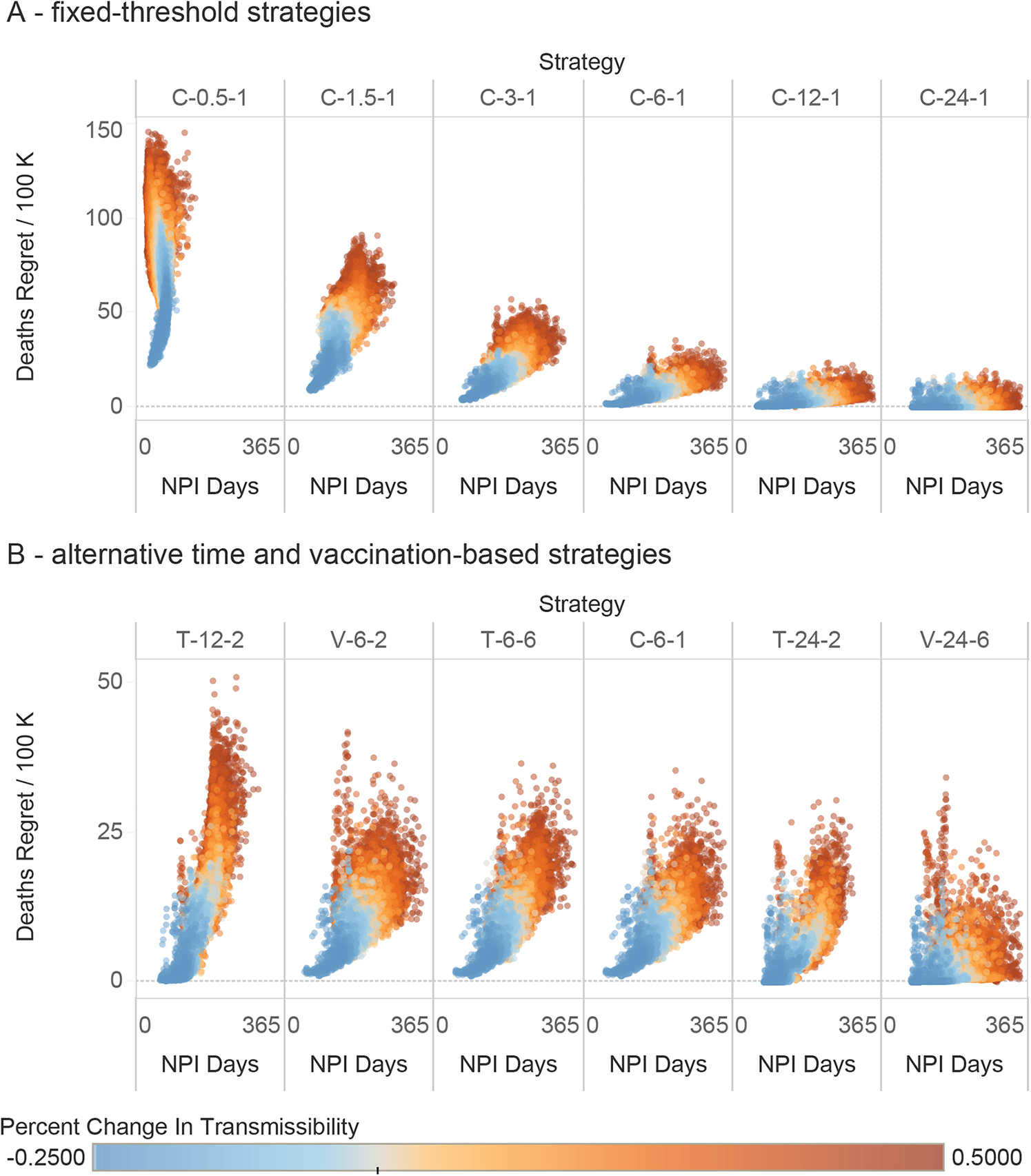
Regret distributions for a sub-set of strategies. This figure presents the performance of a subset of the strategies presented in terms of Deaths / 100 k people regret and Number of Days under NPIs regret. Each dot in this plot represents the performance of each strategy in one of the 20,000 futures. The color gradient represents Change in Transmissibility uncertainty.

Strategy codes are defined as follows. The first two columns describe the characteristics of each strategy. The first letter in the strategy name represents the strategy type (C for constant caution, T for time-based, and V for vaccine-based strategies). The subsequent number represents the baseline level of caution *x*_*b*_, and the third number is a sequential code to make the strategy code unique. The parameters column describes the policy levers that characterize each strategy, as described in the methods section. The final three columns present the 75^*th*^ regret percentile of three metrics of interest.

## Supplemental Information

### 1.1 Model Overview

The model used in this analysis is based on our previously published ODE model [10] and its recent extension [33]. Figure 4 presents an overview of our compartmental model, whereby individuals in our population progress over the different stages of the infection.

**Figure 4:**
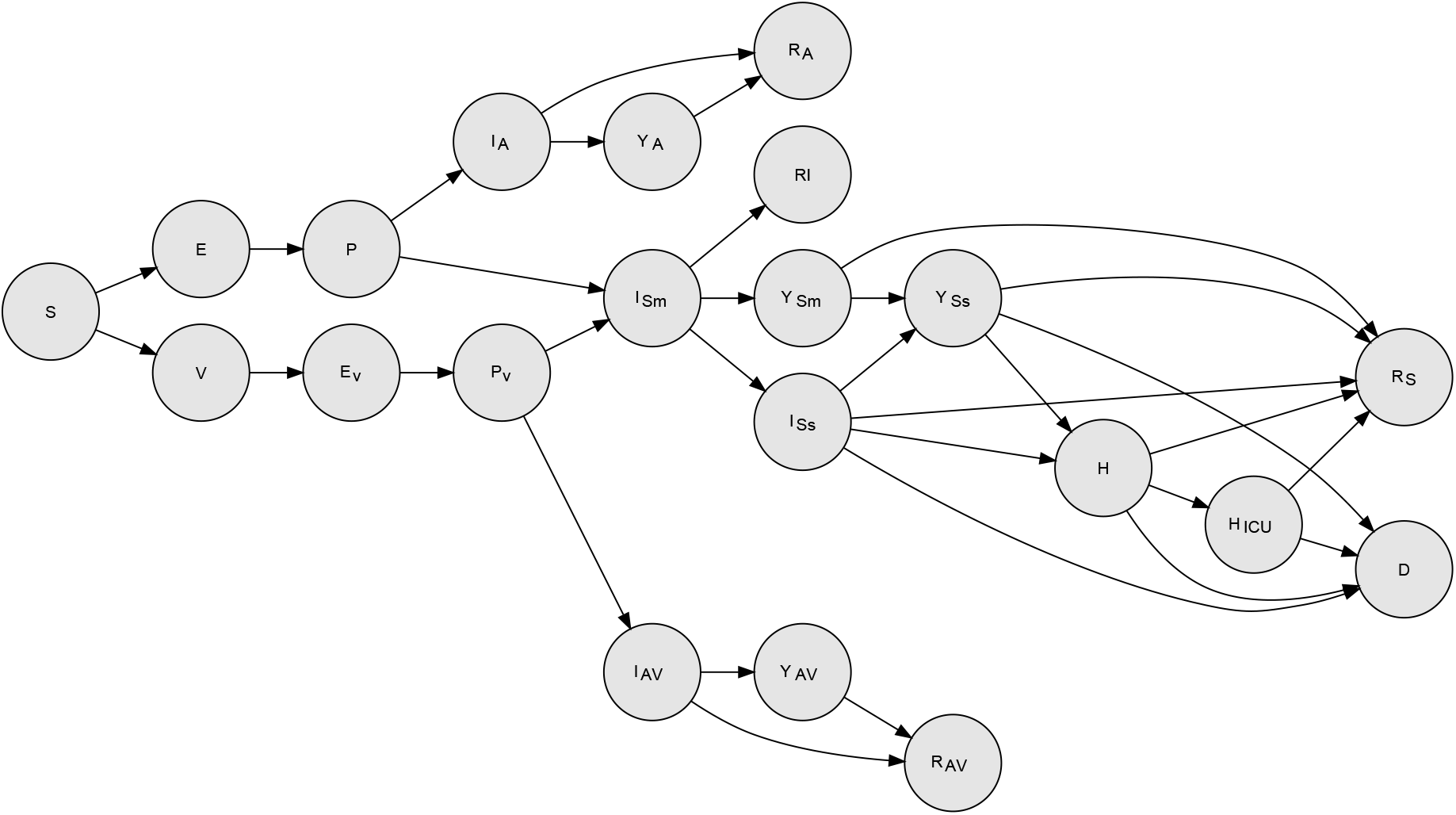
Compartmental Model Structure. Each circle in this figure represents a progression state, each subdivided in 5 population strata. For simplicity and clarity, the figure omits arrows representing loss of immunity (from the recovered states to the S state). See equations for details.

Individuals in our population are divided into 21 compartments. The set of compartments that are common to our NPI-only model include: The noninfected and susceptible (*S*), the exposed and infected but not yet infectious (*E*), the presymptomatic or primary infectious stage (*P*), the infected with mild symptoms (*I*_*Sm*_), the infected with severe symptoms (*I*_*Ss*_), the diagnosed infected with mild symptoms (*Y*_*Sm*_), the diagnosed infected with severe symptoms (*Y*_*Ss*_), the non-ICU hospitalized (*H*), the hospitalized in the ICU (*H*_*ICU*_), the infected asymptomatic (*I*_*A*_), the diagnosed infected asymptomatic (*Y*_*A*_), and those that died (*D*). We assume that individuals in the *P* and *I*_*A*_ compartments are completely asymptomatic and thus are unaware of being infectious. All those compartments were present in our previous work [10, 33].

The model used in this paper includes new compartments aiming to represent vaccination roll-out. Here, we focus our description on these compartments. New compartments include those who have received a full vaccination dose (*V*), the vaccinated who have been infected and are in the exposed and infected but not yet infectious stage (*E*_*v*_), the vaccinated in the presymptomatic infectious stage (*P*_*v*_), the vaccinated in the infected asymptomatic (*I*_*Av*_) and those diagnosed infected asymptomatic (*Y*_*Av*_). This model also has three distinct recovered stages *R*_*I*_, *R*_*A*_ and *R*_*Av*_ allowing us to respectively track those that have recovered having been symptomatic, non-vaccinated asymptomatic, and vaccinated asymptomatic.

The arrows connecting the disease states describe the progression rates between the different compartments. We assume that mild symptoms are a dry cough and a fever, while severe symptoms also include shortness of breath. The sum of the population in all of the states gives the total population *N*. However, we assume that *N* = 1 and thus each state variable gives the proportion of the population belonging to that state.

#### 1.1.1 Model Formulation

Our compartmental model is described by the following set of coupled ordinary differential equations (ODEs):

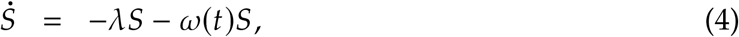

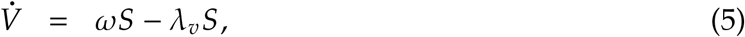

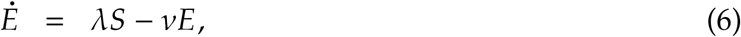

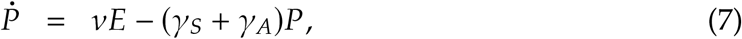

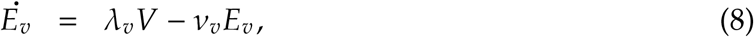

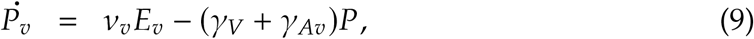

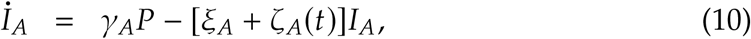

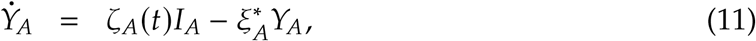

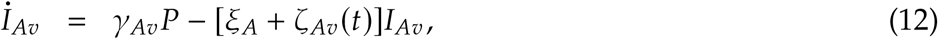

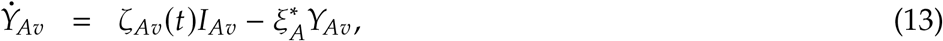

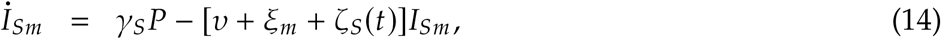

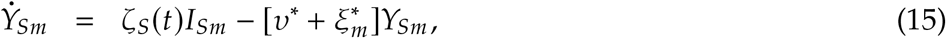

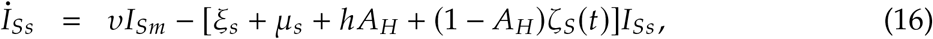

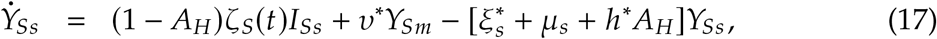

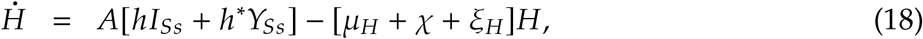

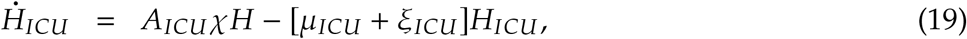

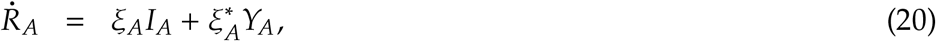

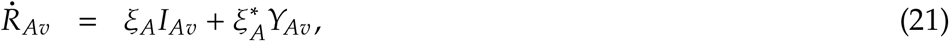

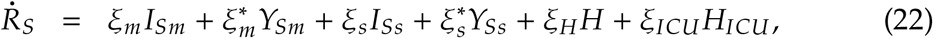

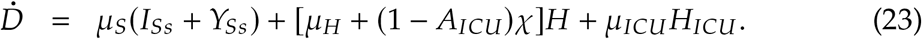

Many of the ODEs and transition rates in equations 4-23 are the same as those used by our first COVID-19 transmission model described in our recent work [33]. Here we focus on providing a high-level overview of the way NPIs are represented in our model and on describing the additions made to the ODEs and the model. Vaccination is the most important addition compared to our previous model, and most of the description is centered around how we model vaccination.

The additional disease compartments include new disease-specific progression rates. In particular, disease progression rates for those that have vaccinate can differ from those who have not. The vaccination rate is given by the parameter *ω* (*t*), described later in this paper. We denote the per-person progression rate from exposure to the presymptomatic for those vaccinated by *v*_*v*_. The progression rates for those who vaccinate differs from the progression rates for those who do not vaccinate. Hence, the progression rates *γ*_*v*_ and *γ*_*Av*_ are not respectively equal to *γ*_*S*_ and *γ*_*A*_. However, we assume that the overall duration of the presymptomatic phase does not change with vaccination. Hence, (*γ*_*v*_ + *γ*_*Av*_)^−1^ = (*γ*_*S*_ + *γ*_*A*_)^−1^. However, the proportion *a*_*v*_ of those vaccinated that remain asymptomatic is higher than the same proportion *a*_0_ of those who did not. Hence, *a*_*v*_ > *a*_0_, where *a*_*v*_ = *γ*_*Av*_(*γ*_*v*_ + *γ*_*Av*_)^−1^ and *a*_0_ = *γ*_*A*_(*γ*_*S*_ + *γ*_*A*_)^−1^. We assume that those who have been vaccinated but get infected and develop mild symptoms progress through the disease’s clinical states as if they were not vaccinated. Hence, for these people, we assume that the vaccine has failed and no longer provides benefits. However, the majority of vaccinated individuals will continue to stay asymptomatic. Their disease progression rate is the same as those asymptomatic who did not vaccinate except for the detection rate. We assume that those who are vaccinated and asymptomatic have a lower rate of seeking to get tested, and hence *ζ*_*Av*_ *< ζ*_*A*_.

Our model also tracks additional outputs. We compute the true cumulative case counts *Ċ*_*T*_; the reported cumulative case counts *Ċ*_*R*_, the cumulative number of people tests 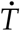, the reported recovered 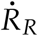, the reported deaths 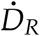 and the reported case-fatality rate CFR_*R*_(*t*). These output quantities are respectively computed using the following equations.

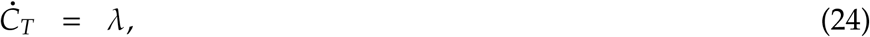

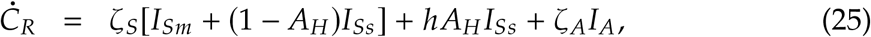

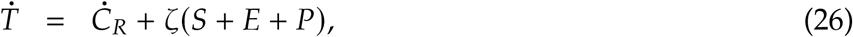

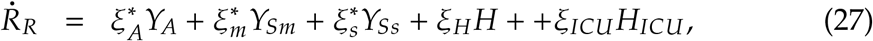

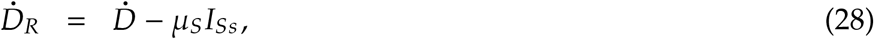

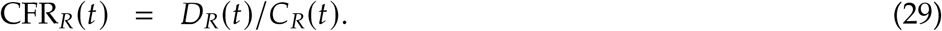

#### 1.1.2 Population Groups and Mixing

Our model considers different population groups or strata. We consider five population strata, including the front-line essential workers (FLEW), the employed non-FLEW, the unemployed, minors of age below 17, and seniors of 65 and above. The first three population strata only include those aged 18 to 64. The prognosis parameters that enter the ODEs depend on the population strata. Prognosis parameters include the proportion of people that develop symptoms (i.e., *γ*_*S*_, *γ*_*A*_, *γ*_*V*_ and *γ*_*Av*_), the proportion of symptomatic who develop severe and critical symptoms (i.e., *v* and *x*), and the proportion of critical cases that lead to death without pharmaceutical treatment (i.e., μ_*ICU*_).

The structure of the model is expressed as an array of ODEs, where the disease progression dynamics for each stratum are expressed by equations 4-23. This formulation extends the model from the more conventional version of a single-strata compartment model that assumes homogeneous mixing and implicit interactions within the population. Heterogeneity in disease transmission is introduced by strata-dependent mixing contact rates describing the variations in how people belonging to the different population strata mix with each other. These strata-dependent mixing contact rates control the transmission dynamics, specifically the *force of infection* terms *λ*. and *λ*._*v*_ that enter the ODEs. We consider six different mixing modes including household, work, school, commercial, recreation, and other. We used a combination of a network-based dataset and self-reported survey data to create matrices describing the average daily contacts between each stratum in each mixing mode [52, 53]. We decompose these matrices into a set of row normalized five-by-five mixing matrices **M**_*m*_, column normalized contact vectors *κ*_*m*_, and scalar mode weight *w*_*m*_ for each mixing mode labeled by the index *m*. The total contact matrix, *K* is calculated by a weighted sum of the mode-specific contact matrices **K**_*m*_, and expressed as

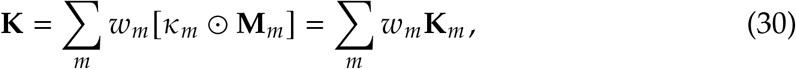

Where ⊙denotes the element-wise multiplication. The weights, *w*_*m*_ give the proportion of contacts (or duration of contacts) of how people mix over the different mixing modes. Under the disease-free status-quo conditions these weights sum to one, hence∑_*m*_ *w*_*m*_ = 1.

#### 1.1.3 Modeling SARS-CoV-2 transmission

SARS-CoV-2 transmission is modeled by the force of infection *λ*. which characterizes how infectious people in each disease state infect others. We express the force of infection as a vector of five elements, one for each stratum and expressed as

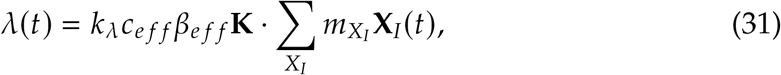

where *c*_*e f f*_ represents the effective contact rate, and β_*e f f*_ effective transmissibility, and *X*_*I*_ represents the set of the infectious disease compartments. This set includes all the disease stages that are infectious, including those that follow from disease transmission of vaccinated people, namely *P*_*v*_, *I*_*Av*_ and *Y*_*Av*_. People who have COVID-19 symptoms or are diagnosed are less likely to mix socially. Moreover, people in the early stages of the disease are more infectious. Hence, we use the coefficients 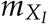 to represents the multiplicative reduction factor for infectious states *X*_*I*_ that scale the transmission rate relative to the asymptomatic and unaware of being infectious.

We compute the value of the product *c*_*eff*_ *β*_*e f f*_ by setting the values of the basic reproductive ratio *R*_0_. By using the next-generation matrix method we find that *c*_*e f f*_ *β*_*e f f*_ = *R*_0_/*t*_*e f f*_, where the time scale *t*_*e f f*_ is expressed in terms of the multiplicative coefficients 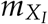 and the values of the disease progression rates [33, 54]. The multiplicative factor *k*_*λ*._represents a calibration constant. Heterogeneity in transmission rates across the population strata is accomplished by the total contact matrix *K*.

Our model assumes imperfect vaccines whereby those who vaccinate may still contract the disease and become infectious and symptomatic. In section 1.1.7 we describe how we model the efficacy of the vaccine and the virus transmission amongst those who vaccinated.

#### 1.1.4 Nonpharmaceutical Intervention Levels

Nonpharmaceutical interventions (NPIs) based on social distancing reduce the total number of unique contacts. They are modeled using a different set of scalar weights *w*_*m*_ that enter equation 30, and are such that their sum is less than one. For example, we can set all values of *w*_*m*_ to zero except for *m* = Household mixing, which retains its original value or perhaps increases it. Additionally, we can modify the mixing matrix **M**_*m*_ for *m* = Household mixing to describing a different behavior of age group mixing within a household due to the new social-distancing measures. Hence, we obtain a different contact matrix *K*. Specifically, to model the impact of reduced mixing from NPI level *n* on mode *m*, we define a diagonal matrix 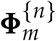. The diagonal elements of 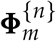 specify the reduction in mixing for each stratum in mode *m* relative to the disease-free state. For interventions that apply to all strata (i.e., where each stratum changes their mixing by the same proportion), such as the closure of schools, all diagonal elements of 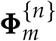 have the same value. However, there are some interventions that only apply to some strata and not others. For example, the case when only essential front-line workers are expected to attend their workplaces. In such cases, the diagonal elements of 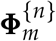 take on different values, each specifying the strata-mode specific impact of the NPI. Hence, the expression for **K**^{*n*}^ that accounts for the impact of NPIs is:

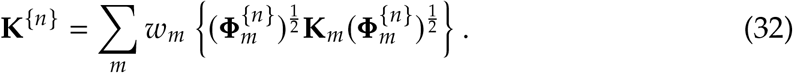

Table 4 provides a description of the intervention levels which are denoted by the index *n*. These intervention levels are used in our model to mechanistically change the transmission processes at different mixing modes and the goal of the model is to compute the consequences of that level of transmission on health outcomes. However, using only those outcomes is not sufficient to properly inform decision-making. As discussed in the methods section, it is desirable to use additional outcome measures to evaluate the pareto-efficiency of alternative strategies. Because minorities and workers at high-contact service industries (e.g., hospitality and leisure) are more likely to face unemployment and income loss during the pandemic [**?**], accounting for the effects of policies on those populations is essential if modelers seek to provide comprehensive decision support to policymakers. For these reasons, we seek to use measures that are monotonically increasing relative to the unknown marginal effect of NPIs on social welfare. This paper uses the number of days of NPIs as the primary measure. In addition to that measure we obtain an estimate of the weekly income loss incurred in each of the NPI levels using the baseline estimates from the general equilibrium economic model [36]. At the end of the simulation run, we aggregate the income loss incurred under each NPI level.

**Table 4:** Nonpharmaceutical intervention levels.. This table describes the NPI levels used by our original model [10]. While the levels could be reorganized to represent alternative preferences (i.e., reopening schools before restaurants), we find that this sequence was roughly equivalent to the sequence used in California. NPI levels implicitly account for adaptation and mitigation measures undertaken to reopen these sectors. Those effects, if observed in the past, are absorbed by an NPI effectiveness factor during calibration.

| NPI Level (*n*) | Description |
| --- | --- |
| Level 1: No Intervention | No Intervention |
| Level 2: Close schools | All schools are closed. |
| Level 3: Close schools, bars, and restaurants; and ban large events | In addition to school closures, all bars' and restaurants' dine-in services are closed, only allowing for take-out options. Also, large gatherings are banned. |
| Level 4: Close schools, bars, and restaurants; ban large events; and close nonessential businesses | In addition to school, bar, and restaurant closures, all nonessential businesses are closed. |
| Level 5: Close schools, bars, and restaurants; ban large events; close nonessential businesses; and shelter in place for the most vulnerable | In addition to the closure of all nonessential businesses, a shelter in place recommended for the vulnerable population, including the elderly, children, and other at-risk populations. |
| Level 6: Close schools, bars, and restaurants; ban large events; close nonessential businesses; and shelter in place for everyone but essential workers | In addition to the interventions above, shelter in place order is issued for everyone but essential workers. |

#### 1.1.5 Modeling Adaptive Strategies

This paper presented only three alternative types of adaptive strategies, which resulted in 78 alternative strategies. Yet, there are many potential ways to frame and model reopening policies. Instead of addressing the question of when society can reopen schools by simulating outcomes under a simple set of rules with an exogenous NPI time-series, we use an endogenous controller to represent a strategy and ask how policymakers should manage their level of caution over time. The difference between the two questions is important. While other studies [8] and our early work [10] addressed the impact of specific fixed policies, this approach does not address the important question of how to adapt policies over time conditional on vaccination. While the first question allows a simple comparison and is more intuitive, the first framing inevitably results in large outbreaks if stringent policies are not followed, and might lead to recommendations that are vulnerable to new strains with higher transmissibility. Because the benefits of NPIs are a non-linear function of the immunity status in the population, and because immunity is changing over time, a set of fixed intervention schedules can result in a menu of options that would be pareto-dominated if a wider set of options was included. This is the main concern and motivation for expanding the option set with alternative strategies.

Framing policies as endogenous also has disadvantages. This formulation implies that policymakers can and will sustain a coherent level of caution over time, and strictly follow that strategy. We remedy this disadvantage by conceptualizing the level of caution as a potentially time-varying control and implementing a stopping condition to cease the use of NPIs once an immunity threshold based on vaccination is crossed. This approach allows us to answer specific questions such as “when interventions can be lifted” while using an endogenous controller that is more robust to uncertainties and representative of adaptive policies. Rather than being an input, the date when NPIs are relaxed is an outcome - a function of policy levers and the uncertainties. The rationale behind this formulation is that policymakers will face higher pressure to relax policies as a wider proportion of the population is vaccinated.

One approach to reconciling the two approaches could be to run the analysis using the endogenous policies over a wide range of futures and then derive an NPI time-series from strategies that were not pareto-dominated. This policy could be translated to an exogenous policy. We did not explicitly do that in this paper because using that NPI time-series for other states or countries could be potentially misleading. However, that approach could be potentially useful for public health departments that wish to translate dynamic, endogenous policies to more interpretable prescriptions.

#### 1.1.6 Vaccination

Our model accounts for a phased vaccination rollout, where a one-dose or a two-dose vaccine is distributed to population strata in order of priority.In our model, those who are immunized (either with a two-dose or a one-dose vaccine) enter the vaccinated compartment *V*. Our model represents vaccination supply and demand separately.

Vaccination capacity is the average rate at which vaccine courses (VCs) can be administered by state. Vaccination capacity increases over time. We assume that, starting from the day when vaccines start to be administered, denoted by *t*_*v*_, the daily supply rate of VCs *s*_*v*_(*t*) increases from zero to a maximum daily rate 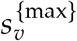 based on the sigmoid function

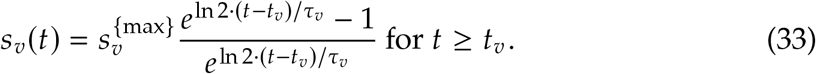

*τ*_*v*_ is the time scale of capacity increase such that 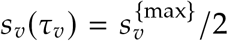. We denote the daily number of VCs utilized in each stratum by the vector **u**(*t*), and the total number of available VCs as a stock variable *v*(*t*). The change in the daily number of available VCs is equal to the difference between the daily number of VCs supplied *s*_*v*_(*t*), and the sum of the daily number of VCs utilized across the population strata, which we denote by *u*(*t*). The latter is equal to the sum of the elements of the vector **u**(*t*). Our model tracks the total number of available VCs *v*(*t*) by treating it as a stock using the following ODE

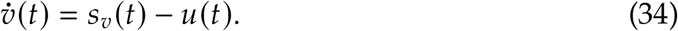

Our model tracks the total number of utilized VCs in each stratum. This is denoted by the vector **U**(*t*) and is given by the time integral of **u**(*t*). The daily number of VCs used **u**(*t*) depends on the vaccination allocation policy and demand. At the start of the vaccine rollout, we assume that policymakers specify a vaccination allocation policy. The policy is denoted by a vector 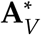. Its elements determine the proportion of vaccines allocated to each population strata, and hence they sum to one. The vector 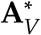 specifies the initial allocation policy, such that higher priority groups have higher values. It is constant over time. However, the actual allocation policy, denoted by **A**_*V*_ (*t*) changes over time because willing members of priority groups deplete as vaccines are distributed. **A**_*V*_ (*t*) depends on the proportion of each stratum willing and eligible to receive additional vaccine doses and the vaccine allocation 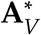.

We define the vector **W** as the proportion of each stratum approved to receive the vaccine and willing to get vaccinated. We then construct an indicator function, ℐ_*V*_ (*t*), which describes if there is still demand in each stratum at time *t*. This allows us to ‘switch off’ vaccine supply to strata that have been fully vaccinated. The indicator function ℐ_*V*_ (*t*) is expressed as a Heaviside step function ℋ (*x*), and compares the number of willing and eligible individuals, **W**, to the total number of utilized VCs, **U**(*t*), element by element:

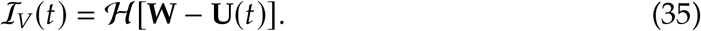

Each element of ℐ_*V*_ (*t*) represents a population stratum. The value of the element is equal to 1 as long as there are still individuals in the stratum approved and willing to vaccinate and equal to 0 otherwise. As of January 2021, the FDA has approved the vaccines for everyone except minors of less than 16. Hence, the value of the ℐ_*V*_ (*t*) for the youngest population strata only considers whether all eligible minors have received the vaccine. The normalized element-wise multiplication of vectors 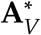 and **D**_*V*_ (*t*), gives the time-varying allocation vector **A**_*V*_ (*t*), and is expressed as

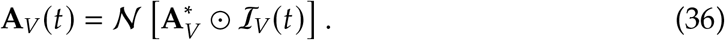

The function 𝒩 (*·*) is a normalization function such that the sum of the elements of **A**_*V*_ (*t*) is equal to one. Thus, as the highest priority stratum has all willing members vaccinated, this value in **A**_*V*_ (*t*) is set to zero, and the priority on other strata are increased.

At the beginning of the rollout, we expect the demand for vaccines to be high. For this case, supply will be limited, and the daily rate of vaccinations in each stratum is given by *s*_*v*_(*t*) · **A**_*V*_ (*t*). However, when the vaccination capacity is no longer a constraint, the daily rate of vaccinations in each stratum no longer depends on the initial vaccination policy 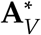. Instead, it depends on demand, which we denote as **D**_*V*_ (*t*). As mentioned, our indicator function ℐ_*V*_ (*t*) signals whether demand is present for each stratum. We assume that demand is limited to the unvaccinated susceptible (*S*), and recovered (*R*_*A*_ and *R*_*S*_) population. Hence, the vector representing the demand for VCs in each population strata is given by an element-wise multiplication of vectors *S* + *R*_*A*_ + *R*_*S*_, and ℐ_*V*_ (*t*), and expressed as

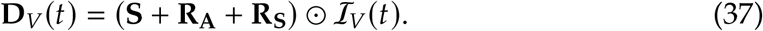

When people no longer perceive the vaccination capacity as constrained, they may seek to get vaccinated at a different rate, *s*_*w*_(*t*). We assume that this probability is the same across the population strata and does not vary with time. We also make the assumption that vaccination rate is independent of the Nonpharmaceutical intervention policy. Hence, in our model the daily consumption rate of VCs is the minimum of supply and demand in each strata:

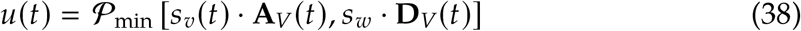

The function 𝒫_min_[**x, y**] is the parallel minimum and returns the element-wise minimum between the vectors **x** and **y**. We can convert this into a per-person daily consumption rate of VCs among the susceptible:

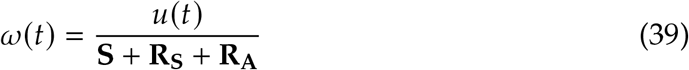

Both the Heaviside step function and the parallel minimum introduce abrupt changes in the model dynamics and lead to a significant increase in stiffness of the ODEs. This is problematic because it significantly slows the numerical solvers. To resolve this issue, we used a continuous approximation to these functions. For example, we approximated the step function with a very steep sigmoid function. For the parallel minimum, we used a “soft” parallel minimum algorithm [**?**].

#### 1.1.7 Vaccination Efficacy

Our model separately considers the vaccine efficacy in protecting from disease transmission and in preventing symptoms. As inputs, the model requires the specification of the vaccine’s overall efficacy of *e*_*v*_ and efficacy in protecting from disease transmission *e*_*tv*_. The overall efficacy is given by

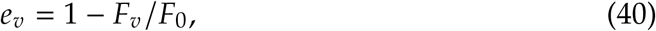

where *F*_*v*_ is the proportion of individuals in the treatment group that during phase 3 vaccine trials reported having symptoms, and *F*_0_ represents the same proportion in the control-placebo group. We can express *F*_*v*_ and *F*_0_ in terms of the *e*_*v*_ as

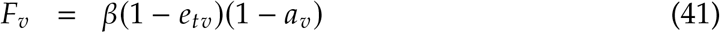

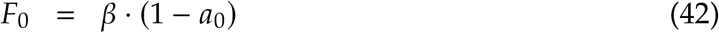

where *β* is the overall transmissibility common to the treatment and the control group. The proportions *a*_*v*_ and *a*_0_ respectively represent the probabilities of remaining asymptomatic after being infected for those who do and do not vaccinate. It follows that we can express *a*_*v*_ in terms of *a*_0_ using the vaccine’s efficacy values by the expression

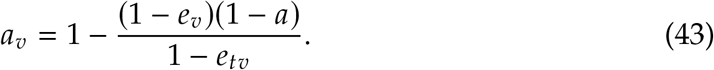

Therefore, by specifying *a, e*_*v*_ and *e*_*tv*_ we can find the value for *a*_*v*_ and hence the relative probabilities that those who are vaccinated develop mild or severe disease, expressed through *γ*_*V*_ and *γ*_*Av*_.

To model the transmission of SARS-CoV-2 to those who have vaccinated we consider both the efficacy of the vaccine in protecting from disease transmission *e*_*tv*_ as well as the increase in the rate of social mixing of those who have vaccinated.

Following from section 1.1.3, we express the force of infection on those who have vaccinated as

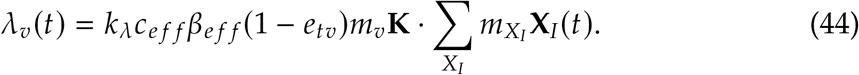

The coefficient *m*_*v*_ represents a multiplicative factor that accounts for the overall effect of behavior changes of the people who vaccinate. For example, these behavioral changes include the tendency for those who vaccinate to be less willing to comply with NPIs and continue to wear their masks, and to generally increase their social mixing rate. In the equations 31 and 44, *X*_*I*_ represents the set of all infectious states and it includes the states *P*_*v*_, *I*_*Av*_ and *Y*_*Av*_. These three infectious states follow from the disease transmission to those who have vaccinated. Hence, both equations depend on the coefficients 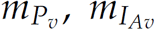 and 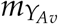 that scale the transmission rate relative to the asymptomatic and unaware of being infectious. The multiplicative factor *m*_*v*_ is included as part of these three coefficients. For example, we set 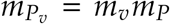, and likewise for the other two factors. Hence, equation 44 considers a squared behavioral effect whereby people who are vaccinated increasingly mix amongst each other by an overall multiplicative factor of 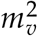.

#### 1.1.8 Additional Mechanisms

Our model includes additional mechanisms that influence the long-term transmission dynamics. These include seasonality and loss of immunity. We model seasonality in transmission by multiplying *c*_*e f f*_ *β*_*e f f*_ by a time-varying term denoted by *ϑ* (*t*) that has an average value equal to one over a year. We use a sinusoidal function to describe *ϑ* (*t*). A parameter *s* controls for the strength of the seasonal effect in the time-varying function *ϑ* (*t*). To model loss of immunity [47], we allow those who recover can become susceptible again. We use a first-order boxcar method and include an intermediate recovered compartment *R*_*B*_, which recovered people transition into before losing immunity and returning the susceptible population pool *S*. These mechanisms are described in more detail in our prior work [33].

### 1.2 Calibration

Model calibration was performed using the Incremental Mixture Approximate Bayesian Computation (IMABC) algorithm [39]. The calibration approach requires the specification of parameter priors *π* (*θ*_*c*_) and calibration targets *y*^∗^. IMABC begins with a rejection-based approximate Bayesian computation (ABC) step, drawing a sample of parameters *θ*_*c*_ of size *N*_0_ from their prior distribution *π* (*θ*_*c*_), simulating calibration targets *y*_*t*_, and accepting parameters that yield simulated outcomes near observed targets within initial tolerance bounds of 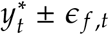. Next, the sample is iteratively updated by drawing additional candidate parameters from a mixture of multivariate normal distributions, centered at the parameters that yield simulated targets that are closest to observed targets. As more points are accepted, the initial tolerance bounds are narrowed, and parameters that yield simulated targets outside of these new bounds are removed. The algorithm has converged when it obtains the requested number of draws that are within final tolerance intervals, 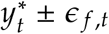. Once the algorithm has converged, posterior estimates can be obtained using a weighted sample from accepted parameter vectors [39]. The weights account for the selection of the sample, using the normal mixture distributions.

We calibrate the model using data from March 1st, 2020 through December 25th, 2020. We use this time period because we are interested in how policymakers should shift their reopening strategy after vaccination started in the US. We use cumulative deaths time-series at the state level, collapsed at ten 30-day intervals *y*_*t*_, *t* ∈ {1, …, 10} starting in March 1st, 2020 through December 25th, 2020. Figure 5(b) shows the data we use and illustrates the model runs that the algorithm selects. We set the initial tolerance level as *ϵ*_*i*_ = *max*(*y*_*t*_) and the final tolerance level as *ϵ* _*f*_ = 0.2*max*(*y*_*t*_) where *y*_*t*_ is the cumulative number of deaths. Therefore, the goal of the algorithm is to find model runs that are within the envelope *y*_*t*_ ±*ϵ* _*f, t*_ of the cumulative death time series illustrated in the graph. In addition to the death time series calibration target, we also require the number of susceptible individuals in the model to be greater than 65%, seeking to find model runs that are consistent with seroprevalence data.

**Figure 5:**
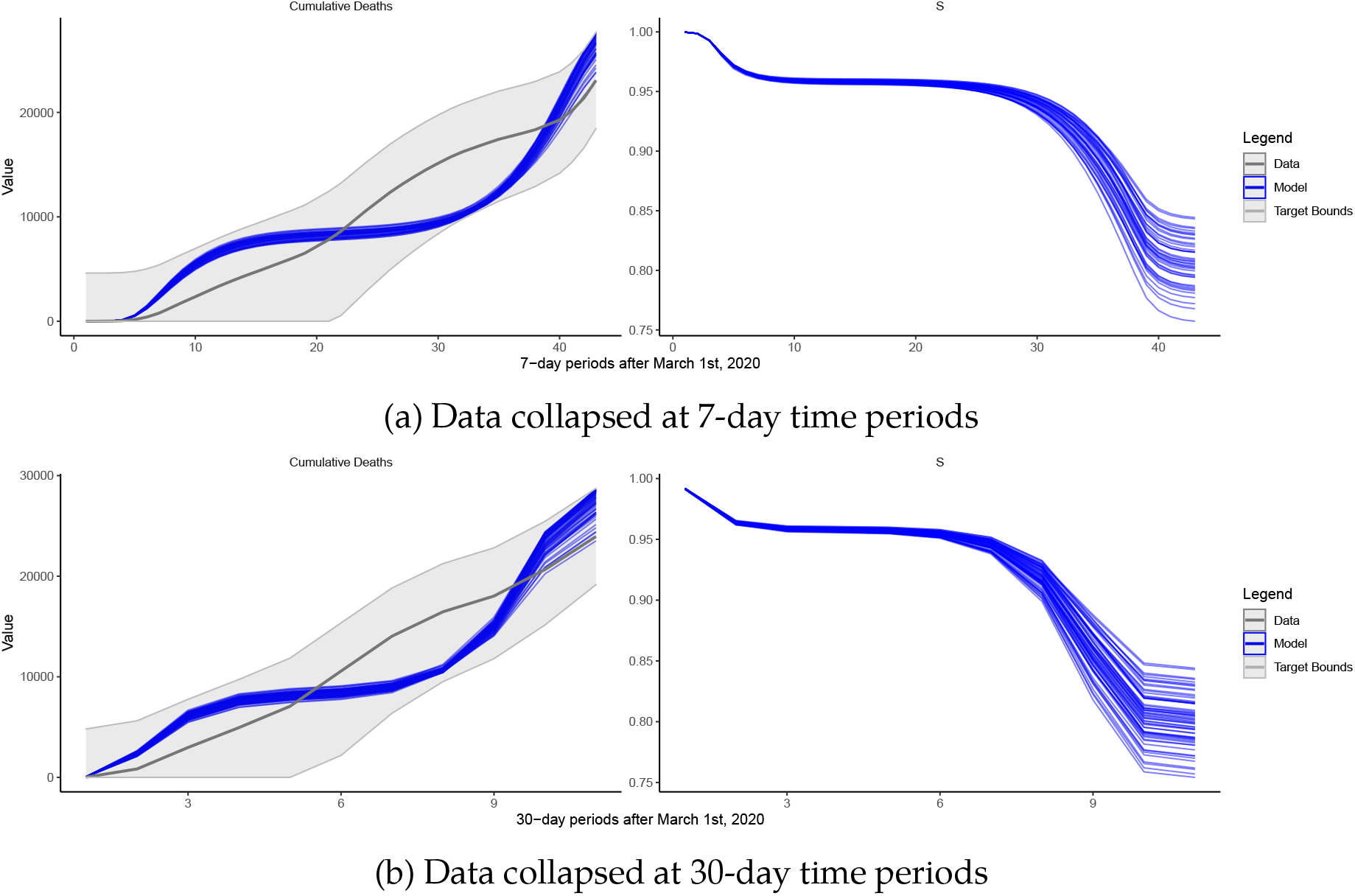
Calibration results for California. The blue lines are the outputs of the calibrated model across three metrics of interest. The data and the calibration target bounds are displayed in gray.

We choose to use 30-day time periods with the aim of reducing the number of individual targets that the calibration procedure needs to track. As figure 5(a) illustrates, choosing another time period (e.g, a 7-day) would yield similar results. Figure 5 also illustrates that the model does not fit the surge in cases around month number 6 in California (September 2020). That is the case because our model does not contain time-varying mixing parameters that could absorb that surge. For the purposes of this analysis, we argue that adding more parameters to the model calibration phase could be problematic^7^.

The calibration results presented above are a function of our model structure presented earlier, the calibration targets and their tolerance interval used, and a set of parameters. Parameters used during the calibration run are divided into three sets: calibrated parameters (C), parameters that were fixed during calibration (F), and parameters that were fixed during calibration but later explored as deep uncertainties (F, D) using a new experimental design. Table 5 presents each parameter, the set to which they belong, a formula or symbol relating the parameter to our model and related sources. The value column includes the parameter mode and minimum and maximum bounds used during calibration.

**Table 5:**
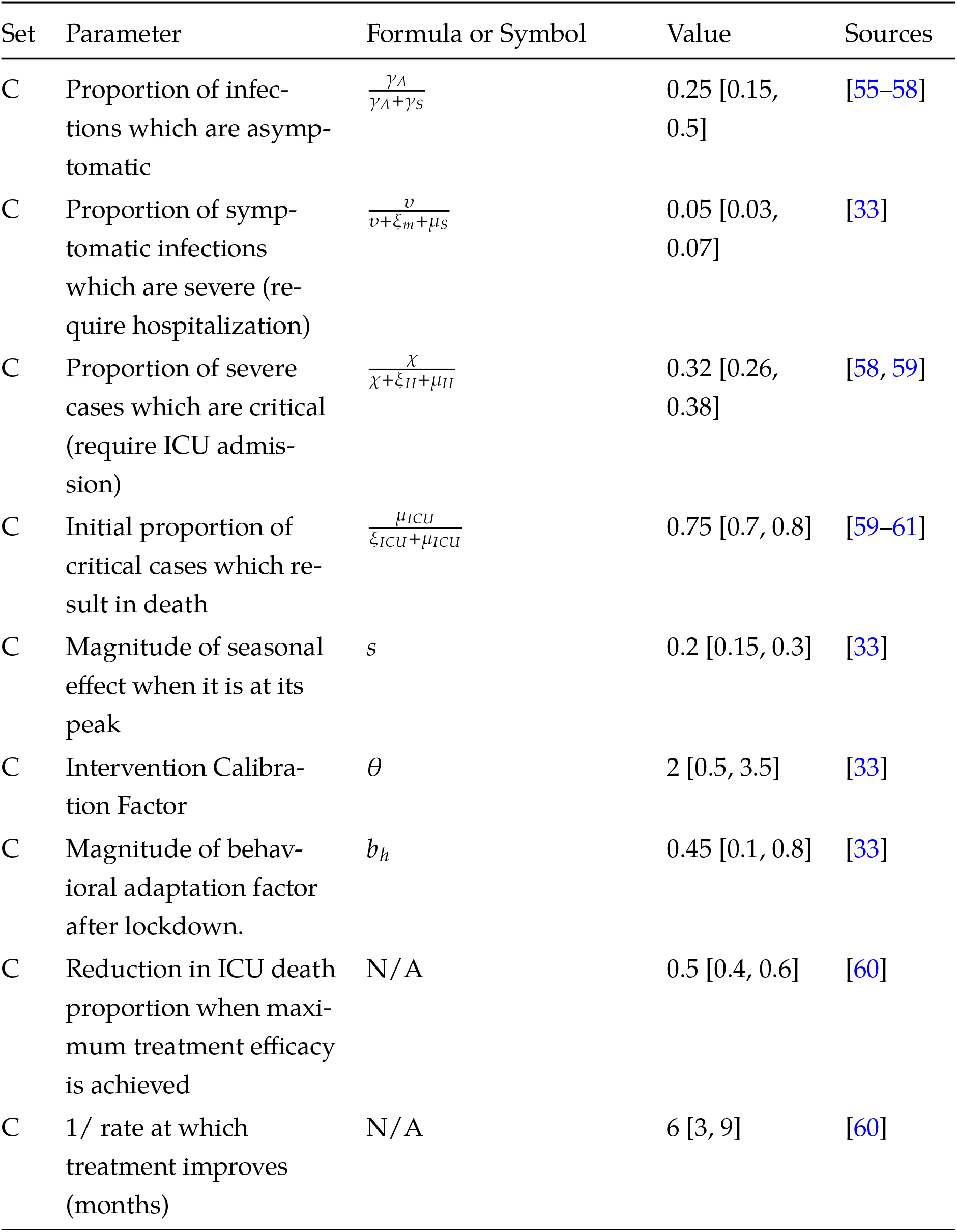
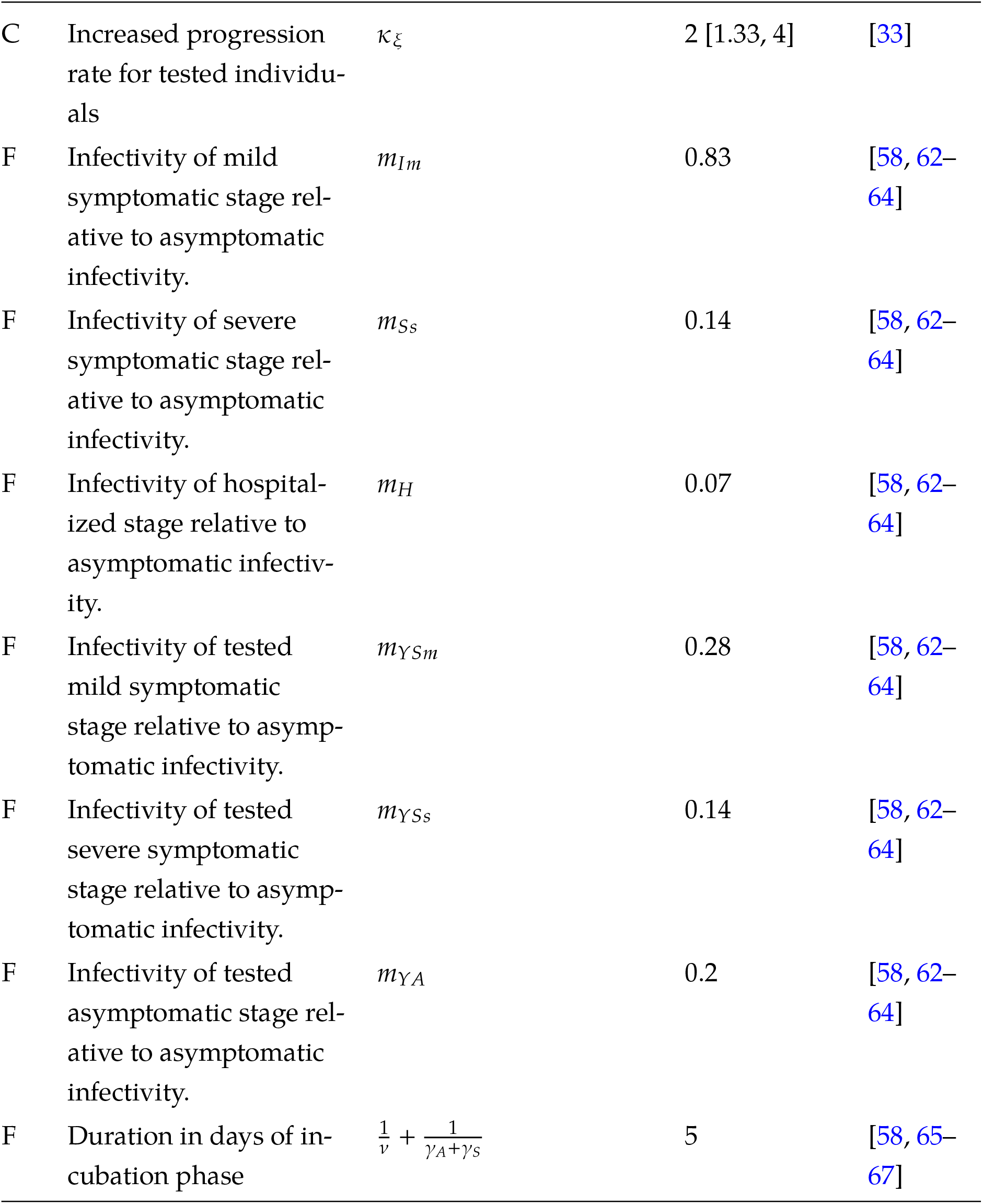
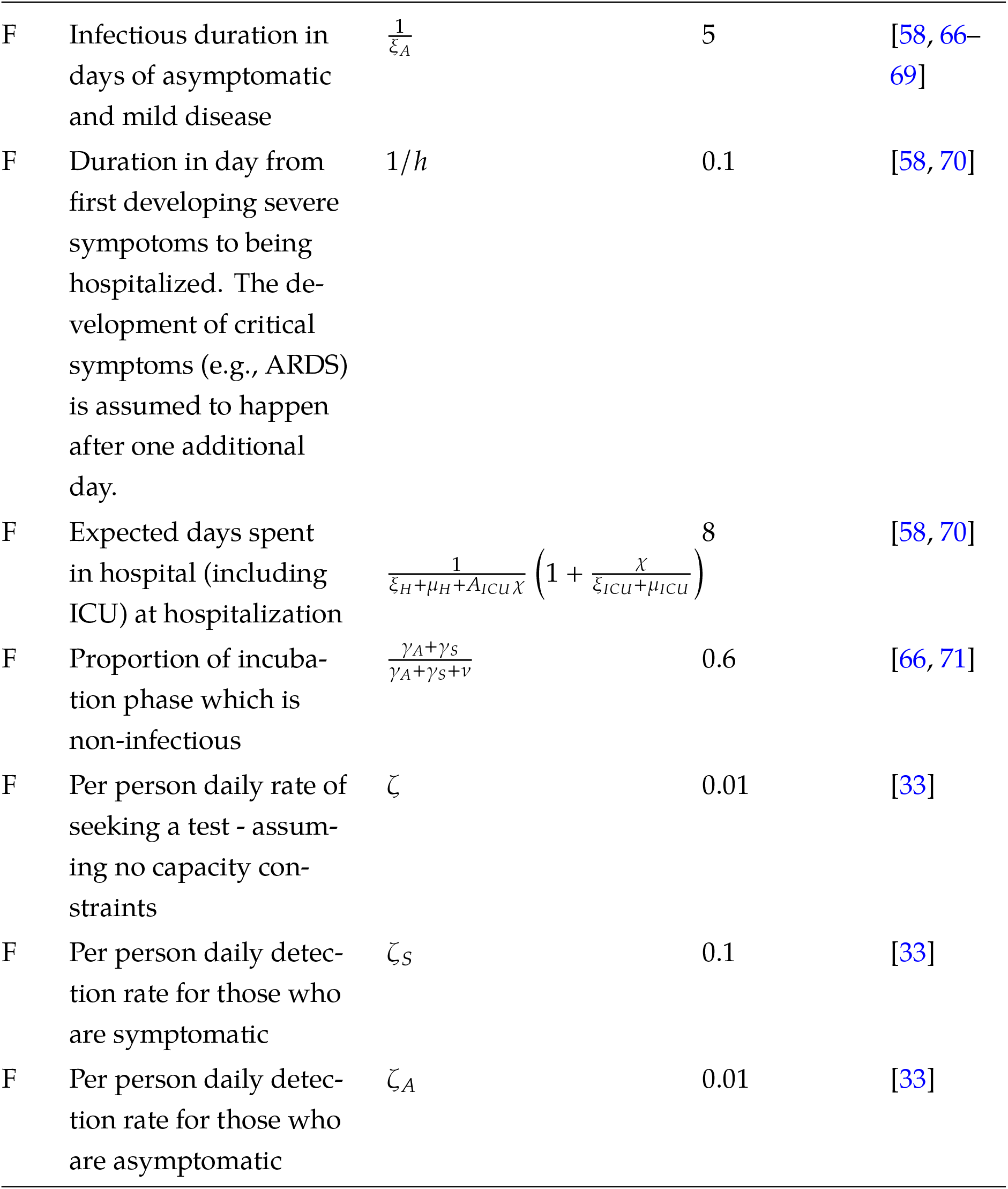
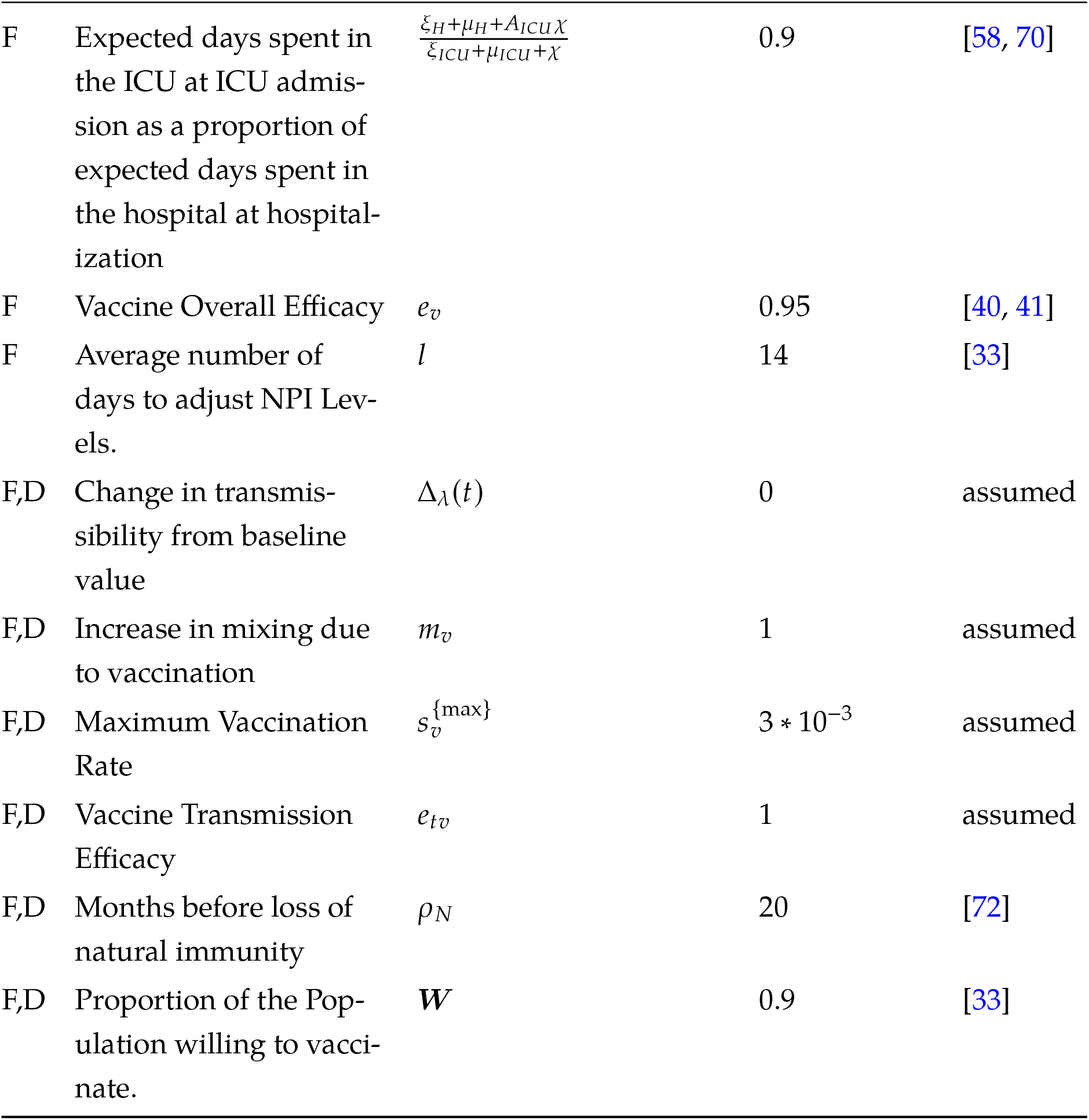
Model Parameters.

| Set | Parameter | Formula or Symbol | Value | Sources |
| --- | --- | --- | --- | --- |
| C | Proportion of infections which are asymptomatic | $\frac{\gamma_A}{\gamma_A + \gamma_S}$ | 0.25 [0.15, 0.5] | [55–58] |
| C | Proportion of symptomatic infections which are severe (require hospitalization) | $\frac{v}{v + \xi_m + \mu_S}$ | 0.05 [0.03, 0.07] | [33] |
| C | Proportion of severe cases which are critical (require ICU admission) | $\frac{\chi}{\chi + \xi_H + \mu_H}$ | 0.32 [0.26, 0.38] | [58, 59] |
| C | Initial proportion of critical cases which result in death | $\frac{\mu_{ICU}}{\xi_{ICU} + \mu_{ICU}}$ | 0.75 [0.7, 0.8] | [59–61] |
| C | Magnitude of seasonal effect when it is at its peak | $s$ | 0.2 [0.15, 0.3] | [33] |
| C | Intervention Calibration Factor | $\theta$ | 2 [0.5, 3.5] | [33] |
| C | Magnitude of behavioral adaptation factor after lockdown. | $b_h$ | 0.45 [0.1, 0.8] | [33] |
| C | Reduction in ICU death proportion when maximum treatment efficacy is achieved | N/A | 0.5 [0.4, 0.6] | [60] |
| C | 1/ rate at which treatment improves (months) | N/A | 6 [3, 9] | [60] |
| C | Increased progression rate for tested individuals | $\kappa_{\xi}$ | 2 [1.33, 4] | [33] |
| F | Infectivity of mild symptomatic stage relative to asymptomatic infectivity. | $m_{Im}$ | 0.83 | [58, 62–64] |
| F | Infectivity of severe symptomatic stage relative to asymptomatic infectivity. | $m_{Ss}$ | 0.14 | [58, 62–64] |
| F | Infectivity of hospitalized stage relative to asymptomatic infectivity. | $m_H$ | 0.07 | [58, 62–64] |
| F | Infectivity of tested mild symptomatic stage relative to asymptomatic infectivity. | $m_{YSm}$ | 0.28 | [58, 62–64] |
| F | Infectivity of tested severe symptomatic stage relative to asymptomatic infectivity. | $m_{YSs}$ | 0.14 | [58, 62–64] |
| F | Infectivity of tested asymptomatic stage relative to asymptomatic infectivity. | $m_{YA}$ | 0.2 | [58, 62–64] |
| F | Duration in days of incubation phase | $\frac{1}{\nu} + \frac{1}{\gamma_A + \gamma_S}$ | 5 | [58, 65–67] |
| F | Infectious duration in days of asymptomatic and mild disease | $\frac{1}{\xi_A}$ | 5 | [58, 66–69] |
| F | Duration in day from first developing severe symptoms to being hospitalized. The development of critical symptoms (e.g., ARDS) is assumed to happen after one additional day. | $1/h$ | 0.1 | [58, 70] |
| F | Expected days spent in hospital (including ICU) at hospitalization | $\frac{1}{\xi_H + \mu_H + A_{ICU}\chi} \left( 1 + \frac{\chi}{\xi_{ICU} + \mu_{ICU}} \right)$ | 8 | [58, 70] |
| F | Proportion of incubation phase which is non-infectious | $\frac{\gamma_A + \gamma_S}{\gamma_A + \gamma_S + \nu}$ | 0.6 | [66, 71] |
| F | Per person daily rate of seeking a test - assuming no capacity constraints | $\zeta$ | 0.01 | [33] |
| F | Per person daily detection rate for those who are symptomatic | $\zeta_S$ | 0.1 | [33] |
| F | Per person daily detection rate for those who are asymptomatic | $\zeta_A$ | 0.01 | [33] |
| F | Expected days spent in the ICU at ICU admission as a proportion of expected days spent in the hospital at hospitalization | $\frac{\xi_H + \mu_H + A_{ICU}\chi}{\xi_{ICU} + \mu_{ICU} + \chi}$ | 0.9 | [58, 70] |
| F | Vaccine Overall Efficacy | $e_v$ | 0.95 | [40, 41] |
| F | Average number of days to adjust NPI Levels. | $l$ | 14 | [33] |
| F,D | Change in transmissibility from baseline value | $\Delta_\lambda(t)$ | 0 | assumed |
| F,D | Increase in mixing due to vaccination | $m_v$ | 1 | assumed |
| F,D | Maximum Vaccination Rate | $s_v^{\{\max\}}$ | $3 * 10^{-3}$ | assumed |
| F,D | Vaccine Transmission Efficacy | $e_{tv}$ | 1 | assumed |
| F,D | Months before loss of natural immunity | $\rho_N$ | 20 | [72] |
| F,D | Proportion of the Population willing to vaccinate. | $W$ | 0.9 | [33] |

Most of our parameters are defined based on clinical evidence. Because disease duration parameters are not informed by our calibration procedure (deaths time-series do not carry information about these rates) and to reduce the dimensionality of the calibration problem, we fix disease duration parameters that can be regarded as less uncertain at this stage in the pandemic. We also fix parameters that define the relative infectiousness of different disease states. Our prior work has established that these parameters have a small influence on the model outcomes we use for calibration when one accounts for the ranges of the other, more uncertain and influential parameters [33].

Other parameters are more uncertain and benefit from calibration. For parameters unique to our model (i.e., the effectiveness of NPIs *θ*) or parameters poorly characterized from existing literature (i.e., magnitude of the seasonal effect on mixing in California during 2020), we use one of two approaches. The first approach is to provide prior ranges and let the calibration algorithm find combinations of parameters that jointly are consistent with the data. The NPI Effectiveness parameter *θ* and the behavioral adaptation factor *b*_*h*_ (to what extent people adapted after the initial lockdowns) are part of this set of parameters. Finally, there are parameters that are regarded as deep uncertainties. Further discussion about how these parameters enter our model is available in our prior work [33]. The next section describes how the full experimental design is created using these parameters.

### 1.3 Experimental Design

Our full experimental design table is composed by the combination of the set of 78 strategies described in the Methods section and listed in Appendix I, the 100 calibration parameters vectors obtained using the calibration approach described earlier and a set of 200 draws from a Latin Hypercube sample of the six deeply uncertain parameters described in table 6. Instead of adding these deep uncertainties to the calibration process, we use baseline values during calibration and explored the uncertainty in their values after calibration. Our approach resembles the scenarios used by the Scenarios Hub initiative [51]. However, instead of defining four discrete scenarios combining uncertainties (e.g. increased transmissibility driven by new variants) and decisions (e.g. relaxation of NPIs), we explore a wider set of strategies under a continuum of plausible futures. Table 6 presents each uncertainty explored in this analysis, the baseline value used during model calibration, and the minimum and maximum values. The baseline value reflects the assumptions commonly made by modelers elsewhere (e.g., there is no endogenous change in transmissibility driven by higher vaccination coverage).

**Table 6:** Deep Uncertainties and their ranges.

| Uncertainty | Symbol | Baseline | Min | Max |
| --- | --- | --- | --- | --- |
| Change in transmissibility from baseline value | $\Delta_\lambda(t)$ | 0 | -25% | 50% |
| Increase in mixing due to vaccination | $m_v$ | 0 | 0 | 20% |
| Maximum Vaccination Rate | $s_v^{\{\max\}}$ | 0.0030 | 0.0023 | 0.0038 |
| Vaccine transmissibility efficacy factor | $e_{tv}$ | 100% | 10% | 100% |
| Average duration of immunity (months) | $\rho_N$ | 20 | 10 | 40 |
| Proportion of the Population willing to vaccinate. | $W$ | 90% | 54% | 90% |

All these uncertainties are applied to the model as an exogenous shift in the level of the baseline uncertainty values after the calibration period. Other functional forms, (e.g., smooth transitions from the baseline value to the new value) could also be implemented to represent these uncertainties with more realism. For example, modeling the impact of variant strains on transmissibility could be done with a logistic growth curve. However, considering these uncertainties using smooth transition functions would require an even larger experimental design to accommodate the additional parameters that control the rate and timing of the changes in parameters, which could be uncertain themselves. To keep our experimental design computationally tractable, we explore uncertainties by changing their values immediately after the calibration phase.

These uncertainties were chosen to include factors often considered by other modelers. For example, at the time of this writing, the MIDAS Network Scenario Hub uses changes in transmissibility driven by variants and vaccination uptake and efficacy [51]. Our uncertainties were chosen to encompass these scenarios and to further explore the impacts of other concerns not commonly addressed in the existing literature. For example, uncertainties in behaviors related to vaccination include the actual maximum vaccination rate that society will achieve, the fraction of the population willing to vaccinate (applied to each population strata), and a potential increase in mixing after vaccination which is often ignored.

When multiple uncertainties are mechanistically entangled in our model (e.g., increases in transmissibility driven by new variant strains can be potentially offset by higher levels of mask-wearing), we use a single parameter that represents the overall change in that parameter from baseline. For example, the change in transmissibility from a baseline value uncertainty Δ_*λ*_ (*t*) is intended to represent the combined effect of more transmissible variant strains [7], increased use of adaptation measures (such as reopening schools with enhanced mitigation protocols), as well as potential changes in the overall mixing. Because all these factors would affect the same transmission equation in our model, sampling multiple uncertainties in a Latin Hypercube to represent separately them would prove inefficient and unnecessary to our purposes. Therefore, we use a single uncertainty parameter to represent the combined effect of these factors.

After calibration, we construct our final experimental design as follows. As described in the calibration section, we obtain a weighted sample from the posterior containing 100 calibration parameter vectors. This sample is obtained with substitution, and resulted in 75 unique parameter sets. We do not need to spend computation time on duplicated parameter vectors. Therefore, we obtain the experimental design by combining the 200 parameter vectors obtained from the uncertainties, with the 75 unique parameter vectors obtained from the calibration procedure. This process results in 15,000 futures under which we test each reopening strategy. We obtain our full experimental design by randomizing the order of the 15,000 futures^8^ and combining this resulting dataset with the set of 78 strategies, which resulted in 1.17 million unique cases to be run. After this process, we re-create the full experimental design by repeating the 25 non-unique calibration parameter vectors according to the number of times they were sampled from the posterior distribution.

The set of decisions we made with respect to our parameters in this particular analysis should not be interpreted as a set-in-stone representation of the pandemic. The set of uncertainties we chose, their bounds and how they were modeled represented our knowledge and concerns with respect to the pandemic in the US as of February 2021. In a regular Robust Decision Making engagement, this set of uncertainties would evolve as more information become available. Parameters that were once deep uncertainties would become regular calibration parameters. For example, if it becomes clear that vaccine prevents transmission, that parameter could be either set during the calibration phase or could be calibrated to data. Similarly, if highly transmissible variant strains become dominant, that change could be reflected in our model with a smooth function. When it becomes clear what will be the final vaccination rate, this parameter could be set. At that point, other uncertainties or policy options might emerge as important, and another analytical cycle could be undertaken to provide further results. The RDM iterative approach would accommodate these new developments, and decision-makers could regularly re-evaluate the robustness of their decisions to the remaining uncertainties of concern.

This section described the additions made to our original model, documented the parameters used in this analysis, and provided details on how the calibration parameters are used with the deeply uncertain parameters and strategies to create our large experimental design that is the basis of our conclusions. Further details about our model can be obtained from our prior work [10, 33].

### 1.4 Computing Environment

The model and the functions used to perform this analysis were implemented in R. The calibration process and the strategy stress-testing runs were performed on Bebop, a High-Performance Computing cluster managed by the Laboratory Computing Resource Center at Argonne National Laboratory. Bebop has 1024 nodes comprised of 672 Intel Broadwell processors with 36 cores per node and 128 GB of RAM and 372 Intel Knights Landing processors with 64 cores per node and 96 GB of RAM. This analysis used slurm’s array jobs to execute the runs in parallel across Broadwell nodes, using 35 cores per node and up to three jobs of 12 nodes at a time.

### 1.5 Code

The model and the functions used to perform this analysis were implemented in R. Readers can find our code and instructions to use it at this github repository [73].

## Footnotes

1 Pareto-dominated strategies are those that are outperformed by another strategy in all criteria under consideration. Because pareto-dominated strategies makes society unnecessarily worse off, they should always be avoided.

2 Some rigorous analyses considered robustness and pareto-efficiency of COVID-19 policies. For example, analyses evaluating vaccination strategies [15, 16] do account for uncertainties and multiple goals.

3 California’s criteria of 7 cases / 100 k to relax the most stringent intervention level translates to a level of caution close to 5 in our model. One can translate a case rate threshold *c* to our model’s level of caution *x* by approximating the known prevalence *p* ≈*c* ∗*τ* from Little’s Law [38], where *τ* is the average duration of the disease and *c* is the case rate criteria, and *p* represents an estimate of the prevalence. From that, and considering our model has five intervention levels, we can translate any case rate criteria to a level of caution by letting *x* = 5 /(*c* ∗*τ* ∗10^3^), which leads to a level of caution of five when *c* = 7 /100*k* and *τ* = 14. We defined the range of the baseline level of caution parameter *x*_*b*_ by exploring the tradeoff surfaces they produce and ensuring that the range includes the edges of the tradeoff space presented in figure 2. We do not include a level of caution of 0 because we regard this level of caution as unrealistic.

4 Alternatively, one might estimate the costs of NPIs using a willingness to pay or a similar approach. As long as the resulting weights are monotonically increasing (i.e., people are not deriving utility from NPI restrictions), our substantive findings would hold because we refrain from aggregating measures. While estimating more precise welfare costs of NPIs and using those costs as a criterion could be valuable to compare benefits and costs at the same scale, we doubt that this approach would lead to precise estimates because these weights are likely not stable over time. Still, as long as these weights are monotonically increasing in the NPI level *at any point in time* our substantive results would hold. Because the weights are highly uncertain and potentially not constant, we refrain from trying to aggregate all outcomes under a single social welfare metric in our analyses as a traditional Cost-Benefit analysis would. Instead, we assess pareto-efficiency and seek strategies that dominate other strategies across a set of outcomes, over a wide range of futures.

5 There are multiple approaches to summarize the many-objective robustness trade-offs implied by alternative strategies when evaluated across a wide range of plausible futures. For example, if one chooses the 100^*th*^ percentile, this approach corresponds to a minimax regret criterion. If one chooses the mean and assumes that future states of the world are equally probable, that would correspond to a Laplace criterion. Here we choose the 75^*th*^ percentile. While changing this percentile can change the strategy rankings, doing so did not change the main substantive conclusions of this paper that constant-caution strategies were dominated.

6 Because all health outcomes are correlated, we use Deaths and Days under NPIs as criteria to perform pareto-sorting and determine which policies are dominated.

7 Adding time-varying parameters to the model seeking to improve model fit without a mechanistic explanation for the summer surge can be problematic for the purposes of this analysis. For example, the surge could signal an increase in pandemic fatigue and not a month-level seasonal phenomenon. Our future work might explore better ways of incorporating realistic behavioral components in our model that could better explain the unexplained surges using more calibration targets to inform the model. For the purposes of this analysis, we chose not to overfit the model to the data and preserve only parameters that represent known mechanisms.

8 This is useful because it allows us to run a fraction of our experiments and evaluate results in the interim, before spending the 50,000 hours of computing time required to run the full experimental design.

## Notes

### Competing Interest Statement

The authors have declared no competing interest.

### Author Declarations

In this modeling study we only used secondary de-identified data.

## References

[1] New York State Government. A Guide to Reopening NY and Building Back Better. Technical Report May, 2020. URL https://www.governor.ny.gov/sites/governor.ny.gov/files/atoms/files/NYForwardReopeningGuide.pdf.

[2] Office of the Governor - Washington State. Safe Start Washington: Phased Reopening County-By-County. Technical report, 2020. URL https://www.governor.wa.gov/sites/default/files/SafeStartPhasedReopening.pdf.

[3] State of California. Blueprint for a Safer Economy, 2020. URL https://web.archive.org/web/20210323005801/https://covid19.ca.gov/safer-economy/.

[4] State of Illinois. Restore Illinois Mitigation plan, 2021. URL https://coronavirus.illinois.gov/s/restore-illinois-mitigation-plan.

[5] Abiel Sebhatu, Karl Wennberg, Stefan Arora-Jonsson, and Staffan I. Lindberg. Explaining the homogeneous diffusion of COVID-19 nonpharmaceutical interventions across heterogeneous countries. Proceedings of the National Academy of Sciences of the United States of America, 117(35):21201–21208, 2020. ISSN 10916490. doi:10.1073/pnas.2010625117. URL https://www.pnas.org/content/117/35/21201.

[6] Marc Lipsitch and Natalie E Dean. Understanding COVID-19 vaccine efficacy. Science, 370(6518):763–765, 11 2020. ISSN 0036-8075. doi:10.1126/science.abe5938. URL https://www.sciencemag.org/lookup/doi/10.1126/scienceabe5938.

[7] Nicolas Castonguay, Wadong Zhang, and Marc-Andre Langlois. Meta-Analysis of the Dynamics of the Emergence of Mutations and Variants of SARS-CoV-2. medRxiv, 2021. doi:10.1101/2021.03.06.21252994. URL https://www.medrxivorg/content/10.1101/2021.03.06.21252994v1.

[8] Katriona Shea, Rebecca K Borchering, William J M Probert, Emily Howerton, LBogich Shouli Li, Willem G Van Panhuis, Cecile Viboud, Ricardo Aguás, Sanjana H Bhargava, Sean Cavany, Joshua C Chang, and Cynthia Chen. COVID-19 reopening strategies at the county level in the face of uncertainty: Multiple Models for Outbreak Decision Support. 2020. doi:10.1101/2020.11.03.20225409.

[9] Stephen M. Kissler, Christine Tedijanto, Edward Goldstein, Yonatan H. Grad, and Marc Lipsitch. Projecting the transmission dynamics of SARS-CoV-2 through the postpandemic period. Science (New York, N.Y.), 368(6493):860–868, 2020. ISSN 10959203. doi:10.1126/science.abb5793. URL https://science.sciencemag.org/content/368/6493/860.full:.

[10] Raffaele Vardavas, Aaron Strong, Jennifer Bouey, Jonathan Welburn, Pedro Nascimento de Lima, Lawrence Baker, Keren Zhu, Michelle Priest, Lynn Hu, and Jeanne Ringel. The Health and Economic Impacts of Nonpharmaceutical Interventions to Address COVID-19: A Decision Support Tool for State and Local Policymakers. 2020. doi:10.7249/tla173-1. URL https://www.rand.org/pubs/tools/TLA173-1.html:

[11] T. Alex Perkins and Guido España. tOptimal Control of the COVID-19 Pandemic with Non-pharmaceutical Interventions. Bulletin of Mathematical Biology, 82(9): 1–24, 2020. ISSN 15229602. doi:10.1007/s11538-020-00795-y. URL https://doi.org/10.1007/s11538-020-00795-y..

[12] Tobias S. Brett and Pejman Rohani. Transmission dynamics reveal the impracticality of COVID-19 herd immunity strategies. Proceedings of the National Academy of Sciences of the United States of America, 117(41):25897–25903, 2020. ISSN 10916490. doi:10.1073/pnas.2008087117. URL https://www.pnas.org/content/117/41/25897.

[13] Jack H Buckner, Gerardo Chowell, and Michael R Springborn. Dynamic prioritization of COVID-19 vaccines when social distancing is limited for essential workers. Proceedings of the National Academy of Sciences, 118(16): e2025786118, 4 2021. ISSN 0027-8424. doi:10.1073/pnas.2025786118. URL https://www.pnas.org/content/118/16/e2025786118.

[14] Molly E Gallagher, Andrew J Sieben, Kristin N Nelson, Alicia N M Kraay, Ben Lopman, Andreas Handel, and Katia Koelle. Considering indirect benefits is critical when evaluating SARS-CoV-2 vaccine candidates. medRxiv : the preprint server for health sciences, page 2020.08.07.20170456, 2020. doi:10.1101/2020.08.07.20170456. URL https://pubmed.ncbi.nlm.nih.gov/32817958/.

[15] Kate M Bubar, Kyle Reinholt, Stephen M Kissler, Marc Lipsitch, Sarah Cobey, Yonatan H Grad, and Daniel B Larremore. Model-informed COVID-19 vaccine prioritization strategies by age and serostatus. Science, 371(6532):916–921, 2 2021. ISSN 0036-8075. doi:10.1126/science.abe6959. URL https://sciencesciencemag.org/content/371/6532/916.full.

[16] Laura Matrajt, Julia Eaton, Tiffany Leung, and Elizabeth R Brown. Vaccine optimization for COVID-19: Who to vaccinate first? Science Advances, 7(6):p eabf1374, 2 2020. ISSN 2375-2548. doi:10.1126/sciadv.abf1374. URL https://advances.sciencemag.org/lookup/doi/10.1126/sciadv.abf1374:.

[17] Nancy McClung, Mary Chamberland, Kathy Kinlaw, Dayna Bowen Matthew, Megan Wallace, Beth P Bell, Grace M Lee, H Keipp Talbot, José R Romero, Sara E Oliver, and Kathleen Dooling. The Advisory Committee on Immunization Practices’ Ethical Principles for Allocating Initial Supplies of COVID-19 Vaccine — United States, 2020. MMWR. Morbidity and Mortality Weekly Report, 69(47): 1782–1786, 11 2020. ISSN 0149-2195. doi:10.15585/mmwr.mm6947e3. URL https://www.cdc.gov/mmwr/volumes/69/wr/mm6949e1.htm.

[18] Sam Moore, Edward M. Hill, Michael J. Tildesley, Louise Dyson, and Matt J. Keeling. Vaccination and Non-Pharmaceutical Interventions: When Can the UK Relax About COVID-19? SSRN Electronic Journal, pages 1–19, 2021. doi:10.2139/ssrn.3753372.

[19] Basma G. Alhogbi, Jay Love, Lindsay T Keegan, Frederick J Angulo, John McLaughlin, Kimberly M Shea, David L Swerdlow, Matthew H Samore, and Damon J A Toth. Continued need for non-pharmaceutical interventions after COVID-19 vaccination in long-term-care facilities. medRxiv, 53(9):p 2021.01.06.21249339, 1 2021. ISSN 1098-6596. doi:10.1101/2021.01.06.21249339. URL https://doi.org/10.1101/2021.01.06.21249339.

[20] Mehul D Patel, Erik Rosenstrom, Julie S Ivy, Maria E Mayorga, Pinar Ke- skinocak, Ross M Boyce, Kristen Hassmiller Lich, Raymond L Smith, Karl T Johnson, and Julie L Swann. The Joint Impact of COVID-19 Vaccination and Non-Pharmaceutical Interventions on Infections, Hospitalizations, and Mortality: An Agent-Based Simulation. medRxiv, page 2020.12.30.20248888, 2021. URL http://medrxiv.org/content/early/2021/01/10/2020.12.3020248888.abstract..

[21] Nicolò Gozzi, Paolo Bajardi, and Nicola Perra. The importance of non-pharmaceutical interventions during the COVID-19 vaccine rollout. medRxiv, page 2021.01.09.21249480,2021. URL http://medrxiv.org/content/early/2021/01/09/2021.01.09.21249480.abstract.

[22] Abba B. Gumel, Enahoro A. Iboi, Calistus N. Ngonghala, and Gideon A. Ngwa. Mathematical assessment of the roles of vaccination and non-pharmaceutical interventions on COVID-19 dynamics: A multigroup modeling approach. medRxiv, 2020. doi:10.1101/2020.12.11.20247916.

[23] Matthew Betti, Nicola Bragazzi, Jane Heffernan, Jude Dvezela Kong, Angie Raad, Teresa Reis, Ricardo Monteiro, Alexandre Afonso, Vitor Pinheiro, Maria Isabel Antunes, Maria Lucilia Araujo, Joao Niza Ribeiro, Anabela Cordeiro-da Silva, Nuno Santarem, Joana Tavares, Alan H Beggs, Vineet Bafna, Alexander Hoischen, Erich D Jarvis, Alka Chaubey, Ravindra Kolhe, and COVID-19 Host Genome Structural Variant Consortium. Integrated Vaccination and Non-Pharmaceutical Interventions based Strategies in Ontario Canada as a Case Study a Mathematical Modeling Study. medRxiv, pages 1–16, 2021. URL https://medrxiv.org/cgi/content/short/2021.01.06.21249272.

[24] CDC. tOperational Strategy for K-12 Schools through Phased Mitigation. Technical report, 2021. URL https://www.cdc.gov/coronavirus/2019-ncov/community/schools-childcare/operation-strategy.html.

[25] Robert J. Lempert, David G. Groves, Steven W Popper, and Steve C Bankes. A General, Analytic Method for Generating Robust Strategies and Narrative Scenarios. Management Science, 52(4):514–528, 4 2006. ISSN 0025-1909. doi:10.1287/mnsc.1050.0472. URL http://pubsonline.informs.org/doi/abs/10.1287/mnsc.1050.0472.

[26] Robert J. Lempert, Steven W. Popper, and Steven C. Bankes. Shaping the Next One Hundred Years: New Methods for Quantitative, Long-Term Policy Analysis. 2003. ISBN 0833034855. doi:10.7249/MR1626. URL https://www.rand.org/pubs/monograph_reports/MR1626.html.

[27] Robert J. Lempert. Robust Decision Making (RDM). In Vincent A. W. J. Marchau, Warren E. Walker, Pieter J. T. M. Bloemen, and Steven W. Popper, editors, Decision Making under Deep Uncertainty, pages 23–51. Springer International Publishing, Cham, 2019. doi:10.1007/978-3-030-05252-2_2. URL http://link.springer.com/10.1007/978-3-030-05252-2_2.

[28] Warren E Walker, Robert J. Lempert, and Jan H Kwakkel. Deep Uncertainty. In Saul I Gass and Michael C Fu, editors, Encyclopedia of Operations Research and Management Science, pages 395–402. Springer US, Boston, MA, 2013. ISBN 978-1-4419-1153-7. doi:10.1007/978-1-4419-1153-7_1140. URL http://dx.doiorg/10.1007/978-1-4419-1153-7_1140.

[29] Robert J. Lempert. Scenarios that illuminate vulnerabilities and robust responses. Climatic Change, 117(4):627–646, 2013. ISSN 01650009. doi:10.1007/s10584-012-0574-6.

[30] Natalie Peyronnin, Mandy Green, Carol Parsons Richards, Alaina Owens, Denise Reed, Joanne Chamberlain, David G. Groves, William K. Rhinehart, and Karim Belhadjali. Louisiana’s 2012 Coastal Master Plan: Overview of a Science-Based and Publicly Informed Decision-Making Process. Journal of Coastal Research, Sp.Issue 6(10062):29–50, 2013. ISSN 0749-0208. doi:10.2112/SI.

[31] Lloyd Dixon, Robert J. Lempert, Tom LaTourrette, and Robert T Reville. The Federal Role in Terrorism Insurance: Evaluating Alternatives in an Uncertain World. Technical Report MG–679, 2007.

[32] David G. Groves, Jordan R. Fischbach, Evan Bloom, Debra Knopman, and Ryan Keefe. Adapting to a Changing Colorado River Making Future Water Deliveries More Reliable Throught Robust Management Strategies. Technical report, 2013. URL https://www.rand.org/pubs/research_reports/RR242.html.

[33] Raffaele Vardavas, Pedro Nascimento de Lima, and Lawrence Baker. Modeling COVID-19 Nonpharmaceutical Interventions: Exploring periodic NPI strategies. RAND Working Paper, 2021. doi:10.7249/WRA1080-1. URL https://www.randorg/pubs/working_papers/WRA1080-1.html.

[34] Serina Chang, Emma Pierson, Pang Wei Koh, Jaline Gerardin, Beth Red- bird, David Grusky, and Jure Leskovec. Mobility network models of COVID-19 explain inequities and inform reopening. Nature, (June), 2020. ISSN 14764687. doi:10.1038/s41586-020-2923-3. URL http://dx.doi.org/10.1038/s41586-020-2923-3.

[35] Jeremy Howard, Austin Huang, Zhiyuan Li, Zeynep Tufekci, Vladimir Zdimal, Helene-Mari van der Westhuizen, Arne von Delft, Amy Price, Lex Fridman, Lei-Han Tang, Viola Tang, Gregory L. Watson, Christina E. Bax, Reshama Shaikh, Frederik Questier, Danny Hernandez, Larry F. Chu, Christina M. Ramirez, and Anne W. Rimoin. An evidence review of face masks against COVID-19. Proceedings of the National Academy of Sciences, 118(4):e2014564118. 1 2021. ISSN 0027-8424. doi:10.1073/pnas.2014564118. URL http://www.pnas.org/lookup/doi/10.1073/pnas.2014564118.

[36] Aaron Strong and Jonathan Welburn. An Estimation of the Economic Costs of Social-Distancing Policies. An Estimation of the Economic Costs of Social-Distancing Policies, (June), 2020. doi:10.7249/wra173-1.

[37] The Atlantic. Racial Data Dashboard, 2020. URL https://covidtracking.com/race/dashboard.

[38] John D. C. Little. A Proof for the Queuing Formula. Operations Research, 9(3): 383–387, 1961.

[39] Carolyn M. Rutter, Jonathan Ozik, Maria Deyoreo, and Nicholson Collier. Microsimulation model calibration using incremental mixture approximate bayesian computation. Annals of Applied Statistics, 13(4):2189–2212, 2019. ISSN 19417330. doi:10.1214/19-AOAS1279.

[40] Inc ModernaTX. International Council for Harmonisation (ICH) Technical Requirements for Registration of Pharmaceuticals for Human Use, E6(R2) Good Clinical Practice (GCP) Guidance. ModernaTX, Inc, 2020.

[41] Fernando P Polack, Stephen J Thomas, Nicholas Kitchin, Judith Absalon, Alejandra Gurtman, Stephen Lockhart,John L Perez, Gonzalo Pérez Marc, Edson D Moreira, Cristiano Zerbini, Ruth Bailey, Kena A Swanson, Satrajit Roychoud- hury, Kenneth Koury, Ping Li, Warren V Kalina, David Cooper, Robert W Frenck, Laura L Hammitt, Özlem Türeci, Haylene Nell, Axel Schaefer, Ser- hat Ünal, Dina B Tresnan, Susan Mather, Philip R Dormitzer, Uğur Şahin, Kathrin U Jansen, William C Gruber, and C4591001 Clinical Trial Group. Safety and Efficacy of the BNT162b2 mRNA Covid-19 Vaccine. The New England journal of medicine, 2020. ISSN 1533-4406. doi:10.1056/NEJMoa2034577. URL http://www.ncbi.nlm.nih.gov/pubmed/33301246.

[42] Austan Goolsbee and Chad Syverson. Fear, lockdown, and diversion: Comparing drivers of pandemic economic decline 2020. Journal of Public Economics, 193: 104311, 1 2021. ISSN 00472727. doi:10.1016/j.jpubeco.2020.104311. URL https://www.sciencedirect.com/science/article/pii/S0047272720301754.

[43] Melissa Diliberti and Julia Kaufman. Will This School Year Be Another Casualty of the Pandemic? Key Findings from the American Educator Panels Fall 2020 COVID-19 Surveys. RAND Corporation, 2020. doi:10.7249/RRA168-4. URL https://www.rand.org/pubs/research_reports/RRA168-4.html:

[44] Casey Mulligan. Deaths of Despair and the Incidence of Excess Mortality in 2020. Technical report, National Bureau of Economic Research, Cambridge, MA, 12 2020. URL http://www.nber.org/papers/w28303.pdf.

[45] Jennie S. Lavine, Ottar N. Bjornstad, and Rustom Antia. Immunological characteristics govern the transition of COVID-19 to endemicity. Science, 371 (6530):741–745, 2 2021. ISSN 0036-8075. doi:10.1126/science.abe6522. URL https://www.sciencemag.org/lookup/doi/10.1126/science.abe6522.

[46] Craig Gundersen, Monica Hake, Adam Dewey, and Emily Engelhard. Food Insecurity during COVID-19. Applied Economic Perspectives and Policy, 43(1): 153–161, 2020. ISSN 20405804. doi:10.1002/aepp.13100.

[47] Christian Holm Hansen, Daniela Michlmayr, Sophie Madeleine Gubbels, Kåre Mølbak, and Steen Ethelberg. Assessment of protection against reinfection with SARS-CoV-2 among 4 million PCR-tested individuals in Denmark in 2020: a population-level observational study. The Lancet, 397, 2021. ISSN 01406736. doi:10.1016/s0140-6736(21)00575-4.

[48] Loïc Berger, Nicolas Berger, Valentina Bosetti, Itzhak Gilboa, Lars Peter Hansen, Christopher Jarvis, Massimo Marinacci, and Richard D. Smith. Rational policymaking during a pandemic. Proceedings of the National Academy of Sciences of the United States of America, 118(4):1–7, 2021. ISSN 10916490. doi:10.1073/pnas.2012704118.

[49] Shou Li Li,Matthew J. Ferrari, Ottar N. Bjørnstad, Michael C. Runge, Christopher J. Fonnesbeck, Michael J. Tildesley, David Pannell, and Katriona Shea. Concurrent assessment of epidemiological and operational uncertainties for optimal outbreak control: Ebola as a case study. Proceedings of the Royal Society B: Biological Sciences, 286(1905), 2019. ISSN 14712954. doi:10.1098/rspb.2019.0774.

[50] Katriona Shea, Michael C. Runge, David Pannell, William J M Probert, Shou-li Li, Michael Tildesley, and Matthew Ferrari. Harnessing multiple models for outbreak management. Science, 368(6491):577–579, 5 2020. ISSN 0036-8075. doi:10.1126/science.abb9934. URL https://www.sciencemag.org/lookup/doi/10.1126/science.abb9934.

[51] MIDAS Network. Announcing the new Scenario Modeling Hub, 2020. URL https://midasnetwork.us/announcing-the-new-scenario-modeling-hub/.

[52] Joël Mossong, Niel Hens, Mark Jit, Philippe Beutels, Kari Auranen, Rafael Mikolajczyk, Marco Massari, Stefania Salmaso, Gianpaolo Scalia Tomba, Jacco Wallinga, Janneke Heijne, Malgorzata Sadkowska-Todys, Magdalena Rosinska, and W. John Edmunds. Social Contacts and Mixing Patterns Relevant to the Spread of Infectious Diseases. PLOS Medicine, 5(3):e74, March 2008. ISSN 1549-1676. doi:10.1371/journal.pmed.0050074.

[53] Kiesha Prem, Alex R. Cook, and Mark Jit. Projecting social contact matrices in 152 countries using contact surveys and demographic data. PLOS Computational Biology, 13(9):e1005697, September 2017. ISSN 1553-7358. doi:10.1371/journal.pcbi.1005697.

[54] P. van den Driessche and James Watmough. Reproduction numbers and subthreshold endemic equilibria for compartmental models of disease transmission. Mathematical Biosciences, 180(1):29–48, November 2002. ISSN 0025-5564. doi:10.1016/S0025-5564(02)00108-6.

[55] Eric A. Meyerowitz, Aaron Richterman, Isaac I. Bogoch, Nicola Low, and Muge Cevik. Towards an accurate and systematic characterisation of persistently asymptomatic infection with SARS-CoV-2. The Lancet Infectious Diseases, 0(0), December 2020. ISSN 1473-3099, 1474–4457. doi:10.1016/S1473-3099(20)30837-9.

[56] Oyungerel Byambasuren, Magnolia Cardona, Katy Bell, Justin Clark, Mary-Louise McLaws, and Paul Glasziou. Estimating the extent of asymptomatic COVID-19 and its potential for community transmission: Systematic review and meta-analysis. Official Journal of the Association of Medical Microbiology and Infectious Disease Canada, 5(4):223–234, December 2020. ISSN 2371-0888. doi:10.3138/jammi-2020-0030.

[57] Diana Buitrago-Garcia, Dianne Egli-Gany, Michel J. Counotte, Stefanie Hossmann, Hira Imeri, Aziz Mert Ipekci, Georgia Salanti, and Nicola Low. Occurrence and transmission potential of asymptomatic and presymptomatic SARS-CoV-2 infections: A living systematic review and meta-analysis. PLoS medicine, 17(9):e1003346, September 2020. ISSN 1549-1676. doi:10.1371/journal.pmed.1003346.

[58] CDC. COVID-19 Pandemic Planning Scenarios. https://www.cdc.gov/coronavirus/2019-ncov/hcp/duration-isolation.html, February 2020.

[59] Semagn Mekonnen Abate, Siraj Ahmed Ali, Bahiru Mantfardo, and Bivash Basu. Rate of Intensive Care Unit admission and outcomes among patients with coronavirus: A systematic review and Meta-analysis. PLOS ONE, 15(7):e0235653, July 2020. ISSN 1932-6203. doi:10.1371/journal.pone.0235653.

[60] Leora I. Horwitz, Simon A. Jones, Robert J. Cerfolio, Fritz Francois, Joseph Greco, Bret Rudy, and Christopher M. Petrilli. Trends in COVID-19 Risk-Adjusted Mortality Rates. Journal of Hospital Medicine, 16(2):90–92, February 2021. ISSN 1553-5606. doi:10.12788/jhm.3552.

[61] R. A. Armstrong, A. D. Kane, and T. M. Cook. Outcomes from intensive care in patients with COVID-19: A systematic review and meta-analysis of observational studies. Anaesthesia, 75(10):1340–1349, October 2020. ISSN 1365-2044. doi:10.1111/anae.15201.

[62] Ashish Goyal, E. Fabian Cardozo-Ojeda, and Joshua T. Schiffer. Potency and timing of antiviral therapy as determinants of duration of SARS CoV-2 shedding and intensity of inflammatory response. medRxiv, page 2020.04.10.20061325, April 2020. doi:10.1101/2020.04.10.20061325.

[63] T. C. Quinn, M. J. Wawer, N. Sewankambo, D. Serwadda, C. Li, F. Wabwire- Mangen, M. O. Meehan, T. Lutalo, and R. H. Gray. Viral load and heterosexual transmission of human immunodeficiency virus type 1. Rakai Project Study Group. The New England Journal of Medicine, 342(13):921–929, March 2000. ISSN 0028-4793. doi:10.1056/NEJM200003303421303.

[64] Xi He, Eric H. Y. Lau, Peng Wu, Xilong Deng, Jian Wang, Xinxin Hao, Yiu Chung Lau, Jessica Y. Wong, Yujuan Guan, Xinghua Tan, Xiaoneng Mo, Yanqing Chen, Baolin Liao, Weilie Chen, Fengyu Hu, Qing Zhang, Mingqiu Zhong, Yanrong Wu, Lingzhai Zhao, Fuchun Zhang, Benjamin J. Cowling, Fang Li, and Gabriel M. Leung. Temporal dynamics in viral shedding and transmissibility of COVID-19. Nature Medicine, 26(5):672–675, May 2020. ISSN 1546-170X. doi:10.1038/s41591-020-0869-5.

[65] Wei-jie Guan, Zheng-yi Ni, Yu Hu, Wen-hua Liang, Chun-quan Ou, Jian-xing He, Lei Liu, Hong Shan, Chun-liang Lei, David S.C. Hui, Bin Du, Lan-juan Li, Guang Zeng, Kwok-Yung Yuen, Ru-chong Chen, Chun-li Tang, Tao Wang, Ping-yan Chen, Jie Xiang, Shi-yue Li, Jin-lin Wang, Zi-jing Liang, Yi-xiang Peng, Li Wei, Yong Liu, Ya-hua Hu, Peng Peng, Jian-ming Wang, Ji-yang Liu, Zhong Chen, Gang Li, Zhi-jian Zheng, Shao-qin Qiu, Jie Luo, Chang-jiang Ye, Shaoyong Zhu, and Nan-shan Zhong. Clinical Characteristics of Coronavirus Disease 2019 in China. New England Journal of Medicine, 382(18):1708–1720, April 2020. ISSN 0028-4793. doi:10.1056/NEJMoa2002032.

[66] Chanu Rhee, Sanjat Kanjilal, Meghan Baker, and Michael Klompas. Duration of SARS-CoV-2 Infectivity: When is it Safe to Discontinue Isolation? Clinical Infectious Diseases: An Official Publication of the Infectious Diseases Society of America, August 2020. ISSN 1058-4838. doi:10.1093/cid/ciaa1249.

[67] Andrew William Byrne, David McEvoy, Aine B Collins, Kevin Hunt, Miriam Casey, Ann Barber, Francis Butler, John Griffin, Elizabeth A Lane, Conor McAloon, Kirsty O’Brien, Patrick Wall, Kieran A Walsh, and Simon J More. Inferred duration of infectious period of SARS-CoV-2: Rapid scoping review and analysis of available evidence for asymptomatic and symptomatic COVID-19 cases. BMJ Open, 10(8), August 2020. ISSN 2044-6055. doi:10.1136/bmjopen-2020-039856.

[68] Min-Chul Kim, Chunguang Cui, Kyeong-Ryeol Shin, Joon-Yong Bae, Oh-Joo Kweon, Mi-Kyung Lee, Seong-Ho Choi, Sun-Young Jung, Man-Seong Park, and Jin-Won Chung. Duration of Culturable SARS-CoV-2 in Hospitalized Patients with Covid-19. New England Journal of Medicine, 0(0):ull, January 2021. ISSN 0028-4793. doi:10.1056/NEJMc2027040.

[69] CDC. Coronavirus Disease 2019 (COVID-19). https://www.cdc.gov/coronavirus/2019-ncov/hcp/planning-scenarios.html, February 2020.

[70] Christel Faes, Steven Abrams, Dominique Van Beckhoven, Geert Meyfroidt, Erika Vlieghe, and Niel Hens. Time between Symptom Onset, Hospitalisation and Recovery or Death: Statistical Analysis of Belgian COVID-19 Patients. International Journal of Environmental Research and Public Health, 17(20), October 2020. ISSN 1661-7827. doi:10.3390/ijerph17207560.

[71] Lirong Zou, Feng Ruan, Mingxing Huang, Lijun Liang, Huitao Huang, Zhongsi Hong, Jianxiang Yu, Min Kang, Yingchao Song, Jinyu Xia, Qianfang Guo, Tie Song, Jianfeng He, Hui-Ling Yen, Malik Peiris, and Jie Wu. SARS-CoV-2 Viral Load in Upper Respiratory Specimens of Infected Patients. New England Journal of Medicine, 382(12):1177–1179, March 2020. ISSN 0028-4793. doi:10.1056/NEJMc2001737.

[72] Jennie S. Lavine, Ottar N. Bjornstad, and Rustom Antia. Immunological characteristics govern the transition of COVID-19 to endemicity. Science (New York, N.Y.), January 2021. ISSN 1095-9203. doi:10.1126/science.abe6522.

[73] Pedro Nascimento de Lima, Raffaele Vardavas, and Lawrence Baker. Github Repository for the paper: Reopening California: Seeking Robust, Non-Dominated COVID-19 Exit strategies, 2021. URL https://github.com/RANDCorporation/covid-19-reopening-california.

